# The longitudinal mediating role of sleep in the bidirectional relationship between depression and functional disability among older adults: A systematic review and conceptual framework

**DOI:** 10.1101/2025.02.27.25323031

**Authors:** Hao Wu, Lu Wang, Hanwu Chen, Wei Gao

## Abstract

**Background:** The increasing prevalence of depression and functional disability in older adults highlights the need for targeted interventions, with sleep as a potentially modifiable factor, yet the longitudinal effects and mediating role of sleep remain poorly understood.

**Methods:** This review and conceptual framework aimed to examine the pairwise bidirectional associations between sleep, depression, and functional disability and identify the longitudinal mediating role of sleep in the bidirectional relationship between depression and functional disability in older adults. The academic databases PsycArticles, PubMed, MEDLINE, Science Citation Index, Social Sciences Citation Index, ProQuest Dissertations and Theses Global, Cochrane, and Scopus were searched for research published in English between January 2000 and June 2024. Systematic review and cohort study designs were eligible. All included studies were assessed for quality using the Critical Appraisal Skill Programme checklist (CASP 2024).

**Results:** 397,289 citations were identified, and 82 studies meeting the inclusion criteria were included. Cohort studies and reviews provide evidence that there is a dynamic reciprocal correlation between sleep, depression, and functional disability in the older population. We propose that sleep may increase the risk of depression and functional disability in the follow-up years, with sleep acting as a potential mediating factor between depression and functional disability. There was a selection bias in the study samples, as most studies focused on specific populations or regions. Moreover, some of the cohort studies included lacked sufficient follow-up time to observe long-term effects.

**Conclusions:** This review and conceptual framework highlight that sleep health can provide crucial insights for mitigating the adverse effects experienced by older adults due to depression and functional disability. For healthcare professionals and policymakers, it provides evidence about prioritizing sleep health as an accessible step to foster a healthy lifestyle.

**PROSPERO registration number:** CRD42024556536.

**What is already known on this topic:** With the increasing aging population, improving the physical and mental health of older adults has become a key social issue. Substantial epidemiological studies have confirmed the existence of bidirectional relationships between depression, sleep disorders, and functional disability in older adults, with all three variables influencing each other. However, the complex interaction mechanisms among these three variables remain unclear, and further research is needed to explore whether sleep plays a longitudinal mediating role between depression and functional disability.

**What this study adds:** This study significantly enhances our understanding by providing robust evidence of the dynamic, bidirectional relationships among sleep, depression, and functional disability in older adults. Unlike previous research that primarily examined pairwise relationships, our study delves deeper by proposing a comprehensive conceptual framework. This framework underscores the potential mediating role of sleep, suggesting that sleep disturbances are not merely consequences of depression and functional disability but also active contributors to their interaction and progression. By elucidating these underlying mechanisms and potential pathways, our study sheds light on the complex interplay among these three variables, ultimately enhancing the quality of life for older adults.

**How this study might affect research, practice or policy:** This study paves the way for deeper investigation into the causal mechanisms connecting sleep, depression, and functional disability. It highlights the critical importance of prioritizing resources for sleep-related research and interventions, recognizing their significant potential to enhance the well-being of an aging population. This holistic approach aims to foster a more comprehensive understanding and effective strategies for promoting healthy aging.

## Introduction

The global population is undergoing a swift aging transition, largely as a result of prolonged life spans and falling fertility rates. Between 2015 and 2050, the proportion of the world’s population over 60 years will nearly double from 12% to 22%.^1^ With the development of an aging society, the trend of depression and functional disability is on the rise in older adults, making it an important public health issue. Functional disability is defined as a diminished capacity or inability to perform basic self-care tasks required for independent living.^2^ All activities of daily living (ADL) and instrumental activities of daily living (IADL) items explained significant variance in quality of life, both physically and mentally.^3,4^ Persons with disabilities have in general poorer health and higher risk of developing secondary health conditions.^5^ Compared with persons without disability, persons with severe disability are more likely to report having depression (over eight times as likely).^6^ Depression is a significant contributor to the global burden of disease.^7^ Approximately 280 million people in the world have depression.^8^ An estimated 3.8% of the population experience depression, including 5% of adults (4% among men and 6% among women), and 5.7% of adults older than 60 years8. In the context of global aging, depression in late life has raised considerable concern because of its relationship to physical illness, functional impairment, and mortality in the elderly population.^9^

Prior epidemiological studies had reported that functional disability and depressive symptoms were closely related, though the relationship might not always be unidirectional,^10–13^ with potential mechanisms suggesting that functional disability can be conceptualized as a stressful condition that may be involved in the onset and progression of depression.^2^ Evidence also suggested that, functional disability as a stressful event increased the likelihood of developing depressive symptoms.^14,15^ As depressive symptoms and functional disability over the life course is dynamic and changeable,^16,17^ it is worth investigating the long-term association between functional disability and depressive symptoms. A substantial body of research had demonstrated a longitudinal bidirectional relationship between depression and functional disability, a phenomenon that persists across different cultural settings and populations.^18–21^

Different health-related factors also play an important role in driving the effects of functional disability and depression among older individuals, with sleep being a modifiable factor that can significantly influence the link to both depression and functional disability. Longitudinal studies had shown that subjective sleep disturbance was a major risk factor for future development of both first-onset and recurrent depressive episodes in older adults.^22–26^ Also, studies showed that there was reciprocal relationship between sleep and depression symptoms.^24–27^ Short sleep duration was associated with an increased risk of onset and recurrent depression, at the same time, depression could shorten the sleep duration of individuals with normal sleep durations.^28^ In addition, a previous longitudinal study showed that older adults with reduced levels of ADL and IADL had a higher risk of having declining sleep quality.^29^ Some research focused on exploring the impact of sleep problems on incident functional disability, with insomnia,^30^ poor sleep quality,^31^ and both excessive and insufficient sleep duration found to increase the risk of functional disability in older adults.^32^ As can be seen, sleep not only has a longitudinal bidirectional relationship with depression but also exert a bidirectional effect on functional disability. Despite there is substantial and clear evidence linking sleep to both depressive symptoms and functional disability, the nature of the interaction among these three and the possible underlying mechanisms remains unclear. Given that sleep health provides clues for understanding the potential causes that could mitigate the negative effects of depression and functional disability, a better understanding of the relationship between sleep, depression, and functional disability is crucial for crafting effective preventive strategies and personalized interventions for older adults.

For the purposes of this review, (1) we aim to testify the pairwise bidirectional associations between sleep, depression and functional disability in older adults. Depression at T1 predicts functional disability at T2, conversely, functional disability at T1 predicts depression at T2. Similarly, depression at T1 predicts sleep problems at T2, conversely, sleep problems at T1 predict depression at T2. Additionally, sleep problems at T1 predict functional disability at T2, conversely, functional disability at T1 predicts sleep problems at T2. (2) We aim to explore the longitudinal mediating role of sleep on the bidirectional relationship between depression and functional disability in older adults. Depression at T1 predicts sleep problems at T2, which in turn predict functional disability at T3. Likewise, functional disability at T1 predicts sleep problems at T2, which then predict depression at T3. We sought to include a wide range of evidence from different regions, rather than following the traditional systematic review process with inclusion criteria, to better understand causal mechanisms and develop a conceptual framework.

## Methods

### Specifying the research question and search strategy

Our research question was “Does a pairwise bidirectional relationship exist between depression, sleep, and functional disability in older adults, and what is the role of sleep in the potential mechanisms linking depression with functional disability?” We proposed to identify evidence across a broad spectrum of exposure, outcomes, and study designs, and to subsequently choose studies that would comprehensively address each facet of the research question, including underlying mechanisms.

The search strategy took a broad approach to capture the full range of relevant literature: terms for depression and functional disability will be combined with terms for sleep. Medical Subject Headings (MeSH) headings or major subject headings will be used for each category, and exploded where possible. Details of the search terms and strategy for each database were provided in **Table S1** (Supplementary Appendix 2).

### Protocol

The protocol was designed in collaboration with experts specializing in mental health and epidemiology. To ensure the originality of our research question, we comprehensively screened several key databases, including the PROSPERO database, the Open Science Framework, and the International Platform of Registered Systematic Review and Meta-analysis Protocols (INPLASY), for any prior registrations or publications. We registered the protocol on PROSPERO. We followed the Preferred Reporting Items for Systematic Reviews and Meta-Analyses (PRISMA) guidelines.^33^ The PRISMA checklist in the **Supplementary Appendix 1**.

### Database searches

Primary electronic search included PsycArticles, PubMed, MEDLINE, Science Citation Index, Social Sciences Citation Index, ProQuest Dissertations and Theses Global, Cochrane, and Scopus. All databases were searched between January 2000 and June 2024.

### Eligibility criteria

We included sources from any country, published between January 2000 and June 2024. Only articles in English will be included. We included studies that explored depression, sleep, and functional disability. Firstly, the studies included must contain at least two main variables: depression and functional disability. We did not specifically set exposure variable and outcome variable. Secondly, we incorporated sleep parameters (e.g., sleep duration, sleep quality, or sleep disorders) as a variable to examine its relationship with depression and functional disability, as well as the potential mechanistic role of sleep.

For studies to be eligible for inclusion, (1) at least half of the participants had to be within the target age range (≥60 years old) or at least half of the participant age-range had to overlap with this age range; (2) at least containing both depression and sleep variables; (3) at least containing both functional disability and sleep variables; (4) at least containing depression, functional disability, and sleep variables; (5) systematic review, prospective and retrospective cohort studies, and longitudinal cohort study designs will be included; (6) only published peer-reviewed journals with full text articles were included.

The exclusion criteria were as follows: (1) articles published before 2000 and not published in English were excluded; (2) articles whose topic was not the variables including depression, sleep, and functional disability were excluded; (3) participants diagnosed with psychological issues other than depression and other samples (children or young people) were excluded; (4) studies that only recruited participants with a specific medical diagnosis (e.g., stroke, diabetes, or other chronic diseases) were excluded; (5) studies that included exposure and outcome variables occurred at the same time were excluded; (6) unpublished articles, papers, conference articles and monographs were excluded.

Studies could assess depression and functional disability using any psychological tests or batteries, self-reported measures, healthcare records, and diagnostic criteria.

Additionally, research can utilize self-reported measures or objective measurements (e.g., polysomnography or wearable devices) to assess sleep parameters.

### Selection process

After searching the databases, the selection of studies were conducted by three researchers, in an independent and blind approach, strictly following the inclusion and exclusion criteria defined in the search protocol. We investigated the title and abstract (title and abstract screening) of the studies to see whether they met our eligibility criteria. If the titles and abstracts were unclear, the full texts were read to avoid the risk of excluding important studies from the systematic review. Any disagreements that may arise were resolved through consensus. Titles and abstracts were independently screened by two reviewers (H.W., L.W.), and conflicts were resolved by consensus. Full texts were independently screened by two reviewers (H.W., L.W.), and discrepancies were resolved through discussion with a third reviewer to reach consensus (H.C.). Studies were duplicated using Endnote and Rayyan. Following this, articles were imported into the online Rayyan data management system for abstract screening (**Figure 1**). To enhance understanding, we conducted a keyword co-occurrence analysis of the included studies (**Figure 2**).

**Figure 1.**
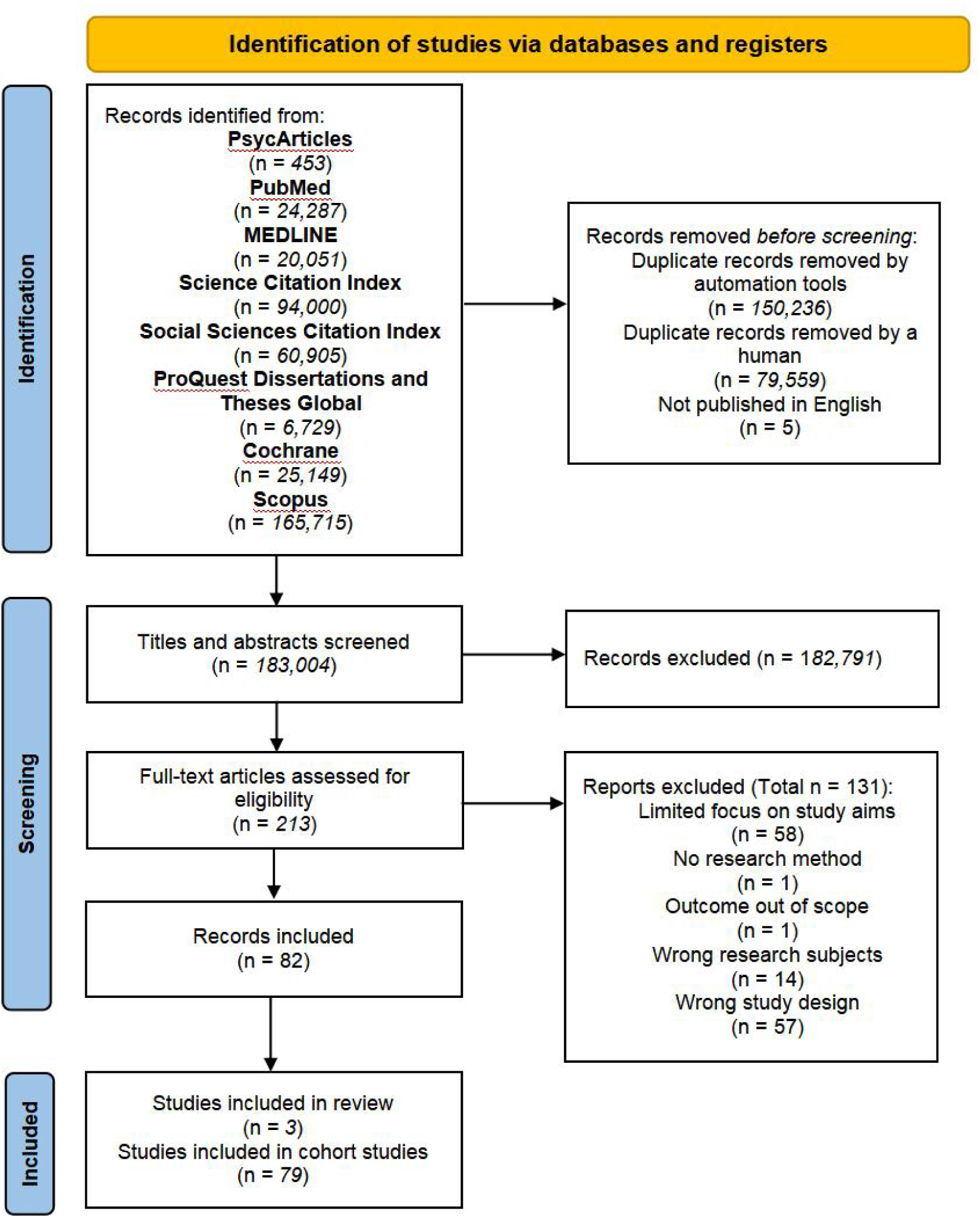
PRISMA flowchart of included studies.

**Figure 2.**
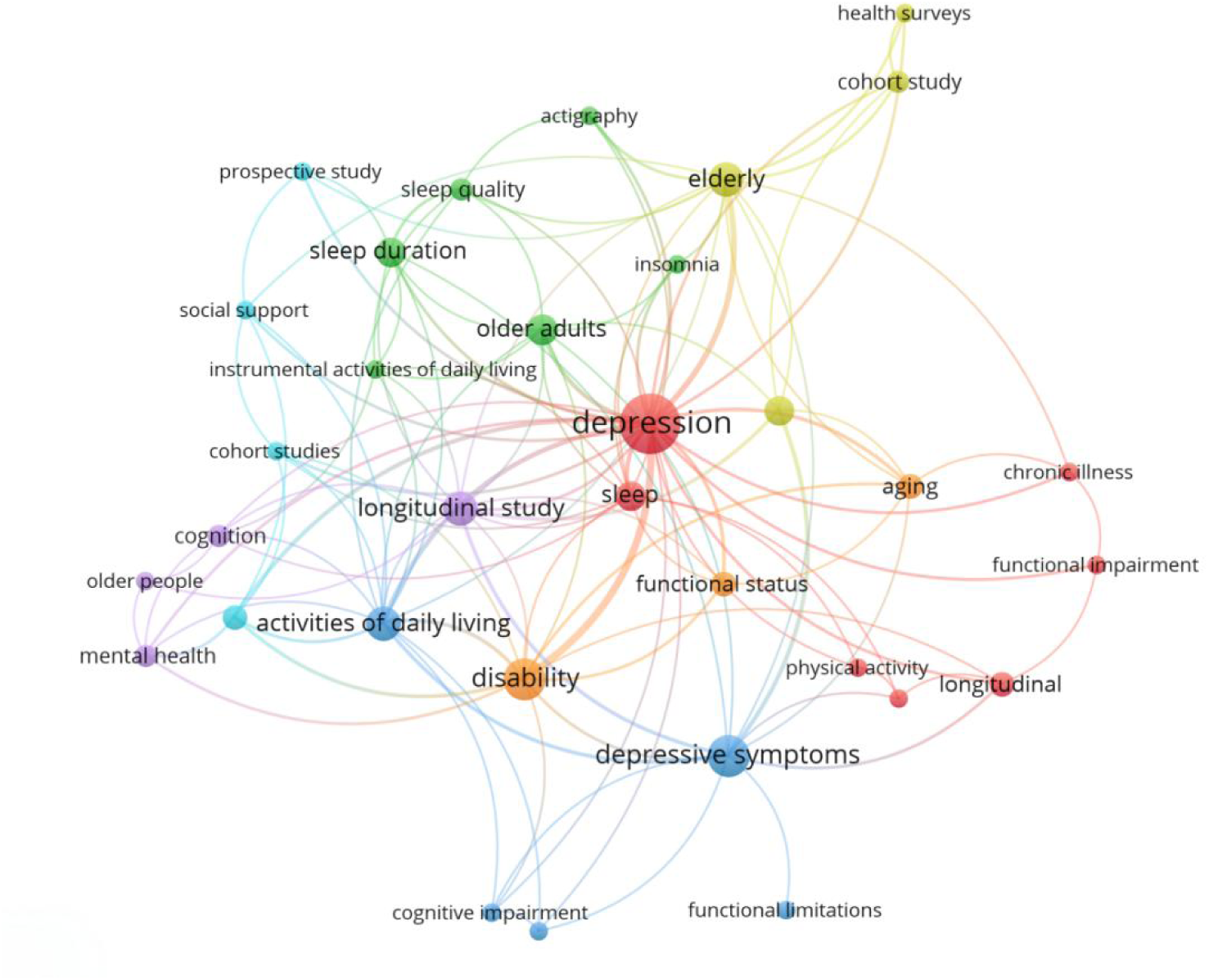
Keyword co-occurrence analysis of the included studies.

### Data extraction and quality assessment

Data extraction carried out independently by three researchers, and any discrepancies will be resolved through discussion. Rayyan will be employed for the storage, management, and retrieval of the data. The following information regarding study characteristics were extracted from the included studies: authors, publication date, study design, aim/objectives/research question, data collection, sample size, exposure measures, outcome measures, follow-up time, covariates, effect sizes, and summary of main findings. When studies reported multiple adjusted models, we extracted information relating to the most adjusted result (the model with the most covariates).^34^ We tabulated the characteristics of each paper, such as exposure, outcome measure, results. Additionally, we recorded information about causal pathways and provided a summary quality score for each paper to facilitate a comprehensive assessment (see **Table S2**, Supplementary Appendix 2). In addition, we summarized the reasons for excluded studies (see **Table S3**, Supplementary Appendix 2).

Quality was assessed using the Critical Appraisal Skills Programme (CASP) checklist.^35–37^ The CASP checklist was used to assess the quality of the articles identified in the searches. The CASP did not have a scoring system to enable studies to be distinctly categorised in terms of quality. Studies were graded in accordance to quality based on the CASP. Studies were graded as followings: ‘low’ (8.5 or higher), ‘medium’ (5-8), or ‘high’ (less than 5) in relation to the risk of having significant methodological flaws.^38^ The primary researcher (H.W.) assessed 100% of the studies. A sample of 20% was independently quality assessed by a second reviewer (L.W.).

Should any disagreements arise regarding inclusion, these were resolved using discussion, or if required, taken to a third reviewer (H.C.), and a majority decision was used to resolve the disagreement (see **Tables S4-S6**, Supplementary Appendix 2). This was in line with the guidance in the Cochrane Handbook for Systematic Reviews of Interventions,^39^ which suggested that a proportion of articles should be reviewed by multiple researchers to check for consistency.

### Framework development

The group members employed a consensus development method for framework development based on formal reviews of the literature. The literature reviews ensured that the framework included all available empirical data. The framework development was an iterative process of drafting, discussions, and revision.^40^ Initially, the framework was built on the basis of strategies for healthy aging, considering the importance of a healthy aging society in promoting the quality of life for the elderly and its long-term developmental benefits. To foster the synchronized development of both the physical and mental health of the elderly, the interconnections between depression, sleep, and physical function were subsequently taken into account. Once the framework was complete, we evaluated the results of the literature review within the context of healthy aging. The conceptual framework is provided in **Figure 3**.

**Figure 3.**
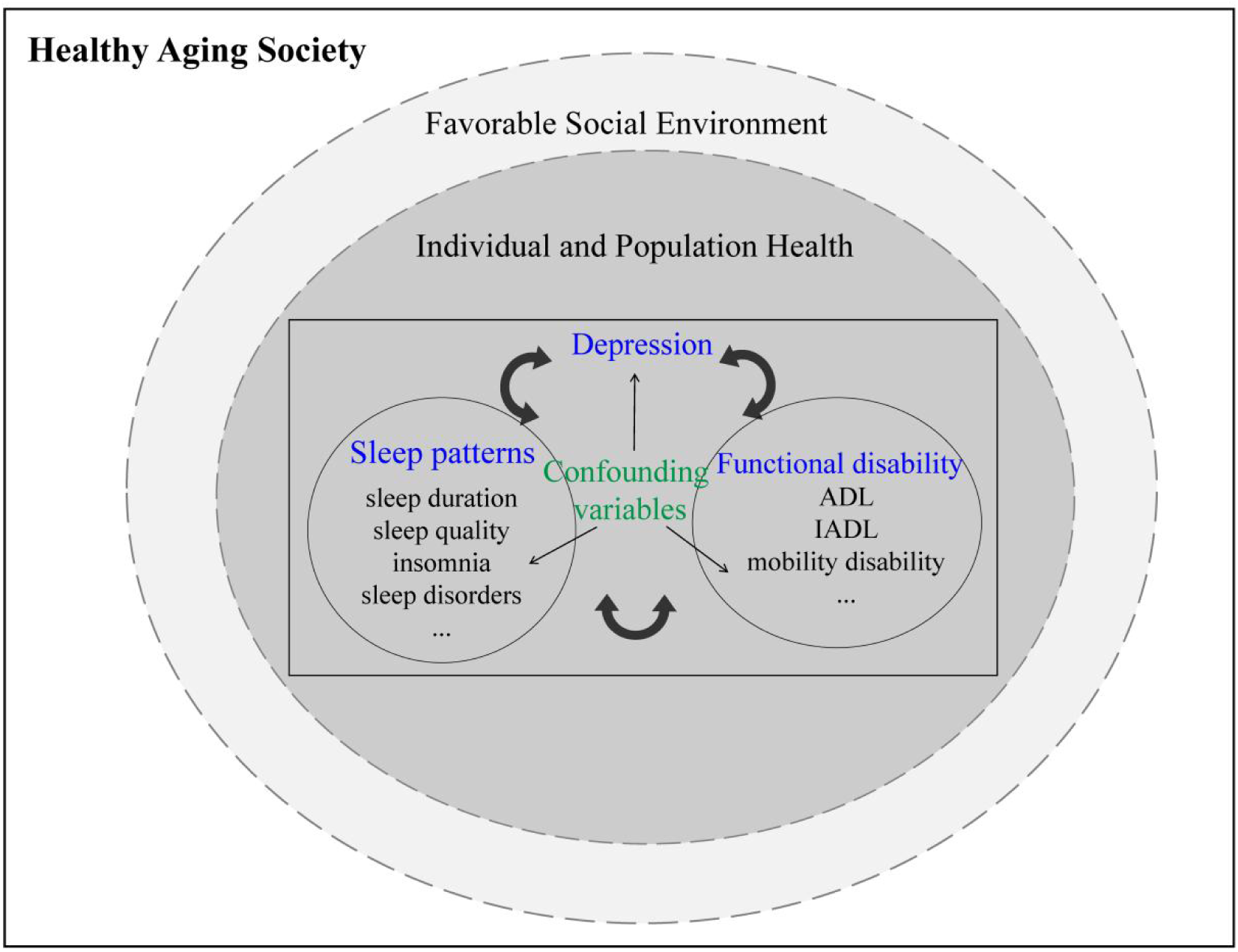
Conceptual framework.

### Synthesis methods and conceptual framework

A narrative synthesis approach will be applied to integrate the findings including an analysis of variability in terms of setting, population, exposure/ intervention, and outcomes.^41^ To comprehensively integrate the research, we revisited the final studies that met the selection criteria. We extracted the key content from each study and compiled it into a tabular format that detailed information on exposure, outcome assessment, results, all aligned with our quality assessment criteria. The main reviewer conducted the preliminary synthesis of each group of studies in regular discussions with other group members. The comprehensive report will give more emphasis to high-quality evidence.

All the information was used to establish two main research hypotheses. The initial conceptual framework aimed to identify the potential mechanisms of depression, sleep, and functional disability in older adults, as well as the factors that may influence this process. We developed two theoretical frameworks to better understand the causal relationships between these three variables. First, we assessed the bidirectional causal relationship between depression, sleep, and functional disability in the older population: depression at T1 predicts sleep at T2, and conversely, sleep at T1 predicts depression at T2; sleep at T1 predicts functional disability at T2, and conversely, functional disability at T1 predicts sleep at T2; depression at T1 predicts functional disability at T2, and conversely, functional disability at T1 predicts depression at T2. Second, we explored the longitudinal mediating effect of sleep between depression and functional disability: depression (T1) predicts sleep (T2), sleep (T2) predict functional disability (T3), and functional disability (T1) predicts sleep (T2), sleep (T2) predict depression (T3). Each part of this framework was reviewed by a group with a psychology background (**Figure 4**).

**Figure 4.**
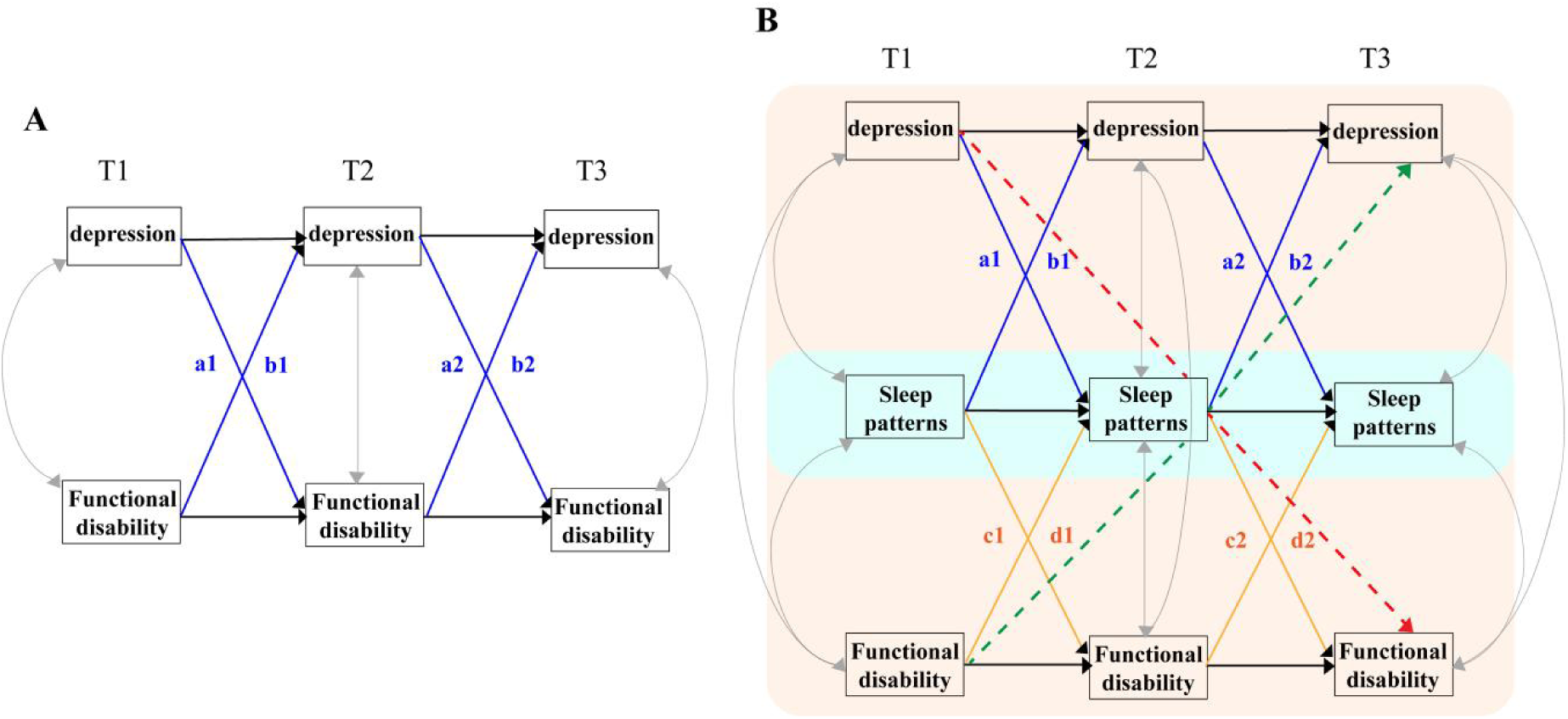
The dynamic relationships between depression, sleep patterns, and functional disability. In Figure A, the black lines represent the autoregressive effects of depression and functional disability; the grey lines indicate the residual correlation between depression and functional disability; a1, a2, b1, b2 denote the cross-lagged effects of depression and functional disability. In Figure B, the black lines represent the autoregressive effects of depression, sleep patterns, and functional disability; the grey lines indicate the residual correlations between depression, sleep patterns, and functional disability; a1, a2, b1, b2 denote the cross-lagged effects of depression and functional disability; c1, c2, d1, d2 represent the cross-lagged effects of sleep patterns and functional disability; the red dashed line indicates the effect of depression on functional disability mediated through sleep patterns; and the green dashed line represents the effect of functional disability on depression mediated through sleep patterns.

## Results

After removal of duplicates, we screened 183,004 titles and abstracts. 213 sources were eligible for full text screening (**Figure 1**). After further exclusions (87.8% of which were due to misalignment with the study aims and wrong study design), 82 sources remained.

### Study design

The majority of studies (n=79, 96.3%) were cohort studies, with only 3 system reviews was included. A wide variation in the duration of included studies, ranging from 10 weeks to up to 20 years. The most common follow-up durations were 4 years (n=16, 19.5%), 2 years (n=11, 13.4%), and 6 years (n=11, 13.4%).

### Study population

Among the 54 cohort studies exploring the longitudinal relationship between depression and functional disability, 68.5% (n=38) of the study populations were aged 60 years and older. Of the 18 cohort studies that assessed the longitudinal relationship between sleep and depression, 33.3% (n=6) included subjects who aged over 60 years. In five cohort studies investigating the longitudinal relationship between sleep and functional disability, one study included a sample of ≥45 years of age, two studies had populations aged 50 years and older, one study focused on population aged over 60 years, and one study selected a population who aged 100 years and older. In two cohort studies examining the longitudinal relationship between sleep, depression, and functional disability, one study was conducted with individuals aged 60 years and older, while the other study included a population starting from the age of 16.

### Study location

The leading countries in the included studies were the United States (n=31, 37.8%), China (n=21, 25.6%), Japan (n=5, 6.1%), and the United Kingdom (n=5, 6.1%).

Among them, 25 (30.5%) studies focused on the relationship between depression and functional disability were from the United States, 13 (15.9%) from China, and 4 (4.9%) from Japan. Of the studies on sleep and depression, there were 5 (6.1%) studies from China and 4 (4.9%) from both the United States and the United Kingdom. Regarding research on sleep and functional disability, 3 (3.7%) studies were from China and one (1.2%) study from the United States. Additionally, there was one (1.2%) study examining the interplay of depression, sleep, and functional disability in older adults from the United States.

### Exposure and outcome exposures and measurements

In research examining the association between depression and functional disability in the older population (n=56), 30 (53.6%) studies designated depression as the exposure variable and functional disability as the outcome variable to assess the long-term effects of depression on functional disability over time. Conversely, 17 (30.3%) studies considered functional disability as the exposure and depression as the outcome to understand the longitudinal impact of functional disability on depressive symptoms. Additionally, 9 (16.1%) studies were conducted to examine the reciprocal longitudinal relationship between depression and functional disability. In studies exploring the relationship between sleep and depression, the included studies all considered sleep as the exposure variable and depression as the outcome variable. Two studies evaluated the bidirectional relationship between the two. The majority of studies are geared towards understanding the prolonged longitudinal effects of sleep on the onset of depression. In the five studies on sleep and functional disability that we included, 4 studies examined the long-term longitudinal impact of sleep as an exposure factor on functional disability, and only one study investigated functional disability as an exposure factor on the long-term longitudinal impact on sleep. Moreover, of the studies exploring the relationship among the three factors, sleep was consistently used as the exposure factor to investigate its impact on depression and functional disability. However, research on the interactive effects among the three and the possible underlying mechanisms had not yet been elucidated.

Most of the assessment for depression were the Center for Epidemiologic Studies Depression (CES-D) scale (n=45, 54.9%), with 9 (11.0%) studies using the Geriatric Depression Scale (GDS), and one of these studies used both the CES-D and GDS scales. Most sleep measurement tools were self-reported (n=17, 20.7%), additionally, four studies used the Pittsburgh Sleep Quality Index (PSQI) to assess sleep quality.

### Quality assessment

After careful evaluation, the majority of included studies were deemed to be of high quality. A total of 11 papers were of medium quality, compared to the high-value papers, the medium-value papers contributed less to the validity of the results and demonstrated limitations in the generalizability of findings. The 11 included studies commonly exhibited limitations, particularly in the presentation of findings, and included: a lack of complete and long-term follow-up of subjects; insufficient control for confounding variables to reduce bias; and the inability to results apply the results to local population. Quality scores were reported in **Table 1**, which presented the outcome of the quality and contribution assessment for each of the 82 included papers.

**Table 1.**
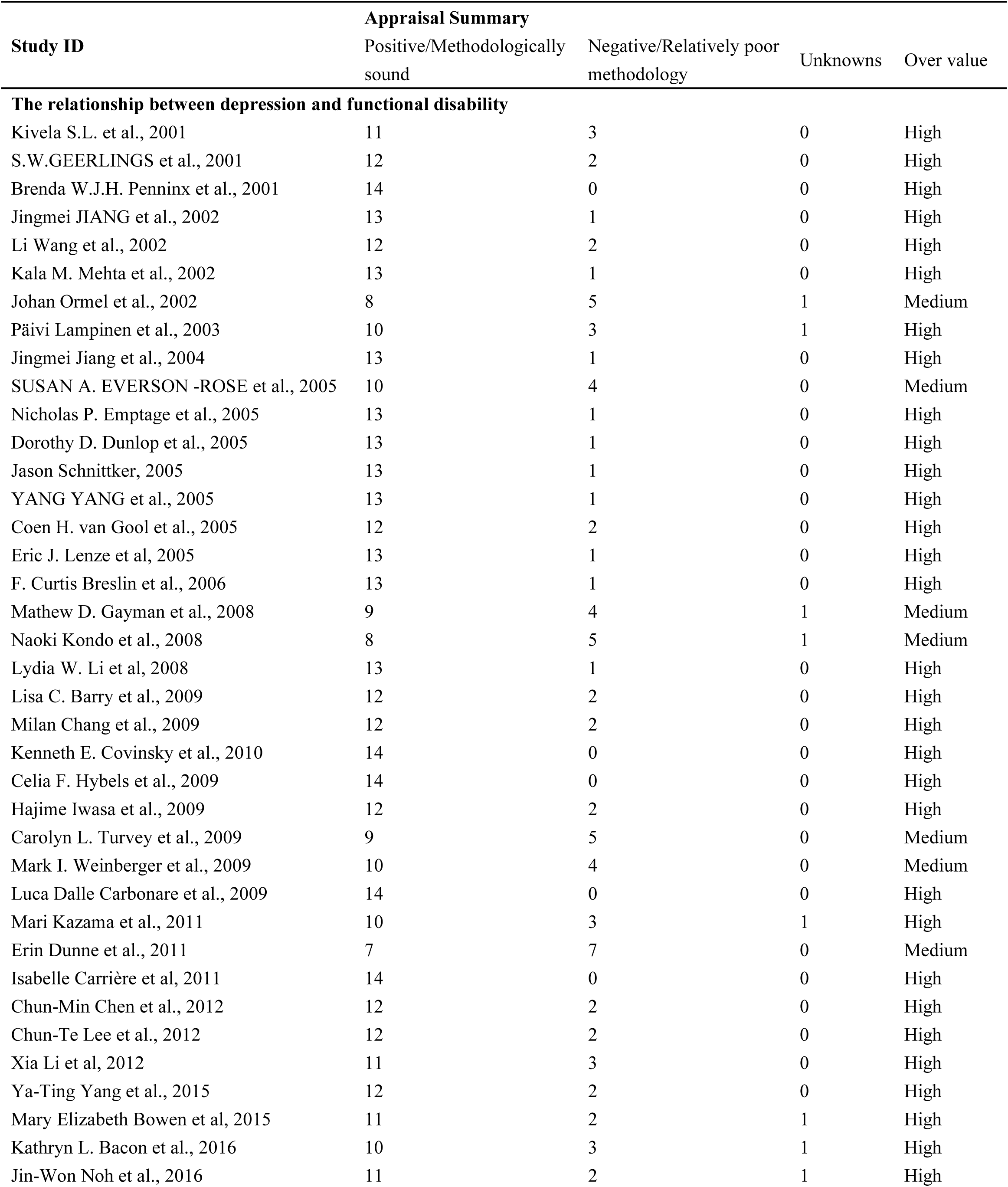
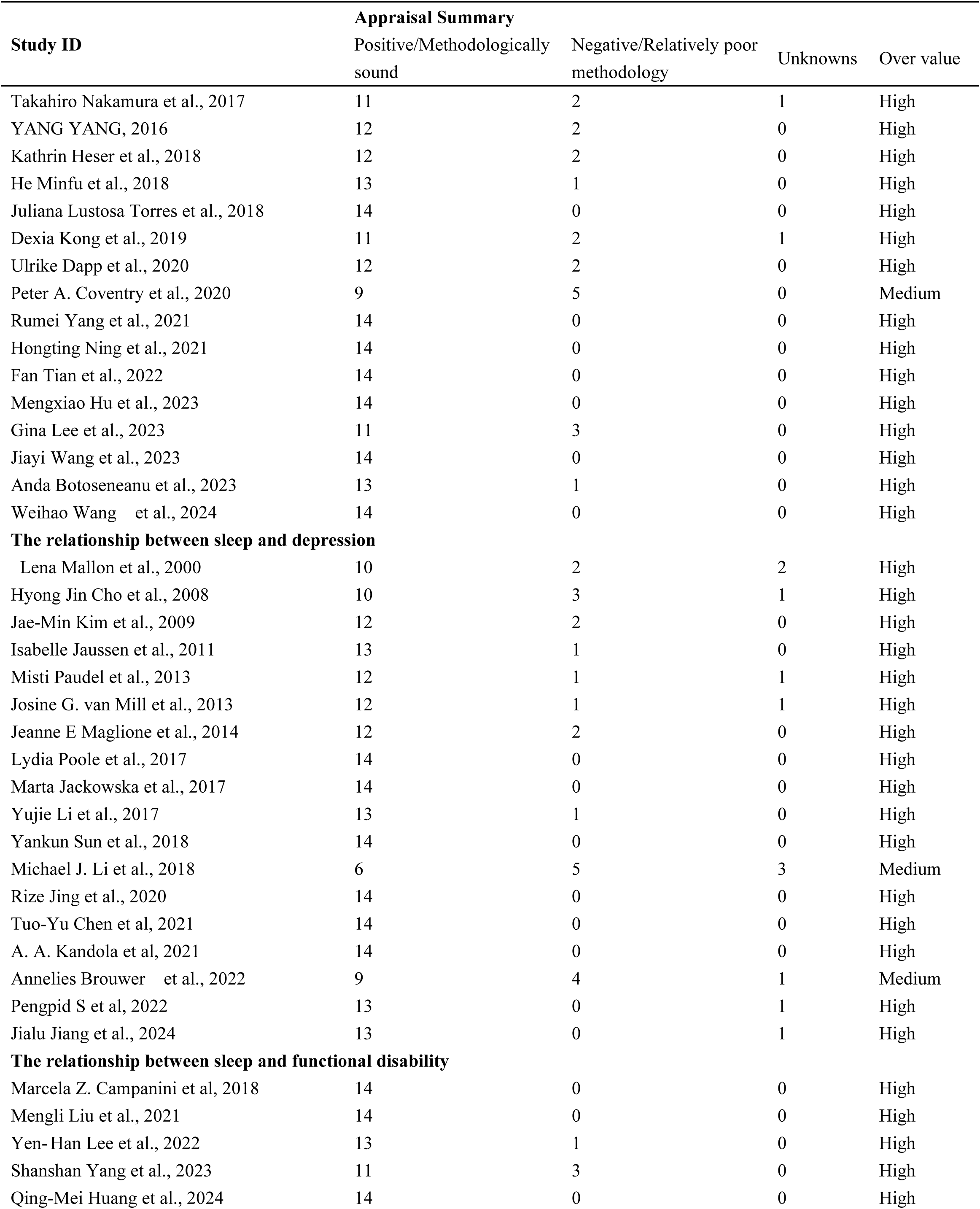
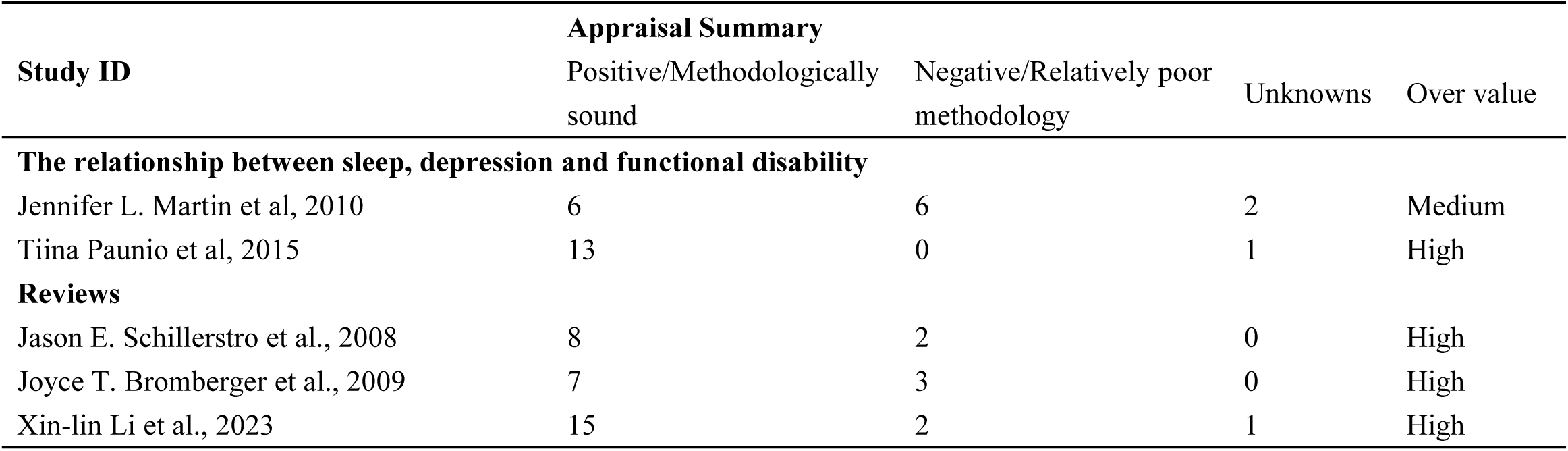
Assessment of methodological limitations.

### The reciprocal relationship between depression and functional disability

Six findings indicated that depressive symptoms were significantly associated with the subsequent disabilities in both activities of daily living (ADL) and instrumental activities of daily living (IADL).^10,42–45^ Among them, one study found that the relative risk associated with depression was stronger for IADL disability than ADL disability ^42^. Seven studies provided evidence that depression remained a strong and significant risk factor for the development of ADL disability,^46–51^ and two studies suggested that those with depressive symptoms were at an increased risk of IADL limitations.^19,52^ Furthermore, one study observed that severe depressive symptoms strongly predicted a subsequent higher IADL decline among the young-old subjects and strongly predicted a subsequent higher ADL decline in the old-old subject.^20^ However, only one study hold the view that depressive symptoms did not consistently contribute to greater physical function decline.^53^ Of the studies investigating the effect of functional disability on the increased depression, three studies testified ADL limitations at baseline were at risk of developing a depressive episode,^54–56^ and two studies considered IADL disability was associated with a higher risk for depression than ADL disability. ^57,58^

Regarding the evaluation of the bidirectional longitudinal relationship between depression and functional disability, one study indicated that a change in state disability had a moderately strong contemporaneous effect on depressive symptoms, whereas a change in depressive symptom level had a weak 1-year lagged effect on disability.^59^ One study compared the effect of the association between depression and functional disability over time, finding that disability was much more likely to affect the increase in the depressive symptom trajectory, as compared with the influence of depressive symptoms on the increase of the disability trajectory over time.^60^ Conversely, another study elucidated that depressed mood predicted subsequent ADL dependency, but ADL dependency did not significantly predict later depressed mood.^61^ In conclusion, there was a correlation between sleep and functional disability in older adults not only within the same temporal dimension, but more importantly, the longitudinal interplay between the two had been widely demonstrated across different study populations and research contexts.

### The reciprocal relationship between sleep and depression

Four studies explored the risk of insomnia leading to an increased risk of depression in older adults,^21,62–64^ while five studies examined the longitudinal relationship between sleep duration and depression.^65–69^ Among them, two studies suggested that both long and short sleep durations were closely related to depression,^65,66^ two studies found that short sleep duration had a greater impact on depression,^28,67^ and one study indicated that long sleep duration had a more significant effect on depression.^68^ In addition, there were inconsistent findings regarding the impact of daytime napping on depression. Some studies suggested that extended napping was associated with a higher risk of depressive symptoms,^69^ however, another research showed that longer midday napping was associated with reduced persistent depressive symptoms.^67^

### The reciprocal relationship between sleep and functional disability

In the five included studies, one study explored that older adults with more ADL and IADL limitations had a higher risk of having declining sleep quality.^29^ One study found that individuals with insomnia symptoms had a higher risk of ADL and IADL disability.^30^ Two studies explored the longitudinal impact of sleep duration on functional disability,^32,70^ with one study showed that long sleep duration increased the risk of ADL disability,^70^ and another study indicated that both long and short sleep durations were linked with incident IADL disability.^32^ In addition, one studies evaluated that poor sleep quality was associated with disability in ADL and IADL.^31^

### The interplay between sleep, depression, and functional disability

Two included studies indicated that poor sleep was associated with declining functional status and greater depression in older adults over the follow-up period.^71,72^

### The conceptual framework

We developed a conceptual framework, based on the reviewed literature, that connects sleep to both depression and functional disability, thereby extending existing research (**Figure 3**). The model incorporated evidence about the longitudinal interplay between sleep, depression, and functional disability in older adults. The framework demonstrated a longitudinal bidirectional lagged relationship between depression and functional disability, such that depression at T1 not only affected functional disability at T1 but also influenced functional disability at T2. In addition, functional disability at T1 not only impacted depression at the same time but also exerted an effect on depression at T2. To explore the potential mechanisms between the two, our framework also suggested the longitudinal mediating role of sleep, where depression at T1 could influence functional disability at T3 through sleep at T2, and conversely.

### Analysis by subgroup

Few studies compared results for males and females, some studies only focused on the interests of males or females. About the relationship between depressive symptoms and functional disability, one study found that mild depressive symptoms was associated with increased incident limitations on IADL in men, whereas severe depressive symptoms was related to IADL and ADL limitations in women.^73^ The impact of depression on increased IADL limitations was greater in males than in females, while the impact of depression on ADL limitations was the most significant in females. What’s more, two studies found that females with worsening disability were more likely to develop depressive symptoms. Research indicated that depressive symptoms and disability formed reciprocal longitudinal relationships in older women.^74,75^ In contrast, one study showed that depressive symptoms predicted a greater risk of subsequent disability in males than in females. Additionally, the relationship between ADL disability and increased depressive symptoms was stronger in females than in males.^21^

About the relationship between sleep and depressive symptoms, two studies suggested that poor sleep quality and long sleep duration were associated with increased odds of depression several years later among men,^76,77^ while another two studies considered that sleep disorders and insomnia were risk factors for susequent depression in women.^78,79^

## Discussion

We synthesized a wide range of evidence about the longitudinal reciprocal relationships between sleep, depression, and functional disability among older adults, and developed a conceptual framework linking the longitudinal mediating role of sleep in preventing depression and functional disability. Our conceptual framework emphasized that sleep health played a pivotal role in the complex interplay between depression and functional disability among older adults. It highlighted the need to prioritize sleep health as a key intervention target to mitigate the adverse effects of these conditions and promote overall well-being in older populations.

Overall, the study results were consistent, with only two studies presenting contradictory conclusions.^53,80^ Nonetheless, the included studies collectively supported our research hypotheses. Firstly, research on depression and functional disability among older adults tended to focus on depression as a primary factor affecting functional disability. The majority of studies measured depressive symptoms using self-reported scales, indicating that depression identified through scales, in addition to clinical diagnoses, warranted serious consideration. Clinical proved that it was very difficult to completely cure severe depression, so early diagnosis and early treatment to avoid exacerbations over time was the most effective way to deal with depression.^81^ Existing studies had already demonstrated that both short-term and long-term presence of depression could severely impact functional disability of older adults.^20,46^ Moreover, baseline functional disability also negatively influenced subsequent depression in older adults. Options varied on whether ADL disability or IADL disability had a greater influence on the incidence or recurrence of depression. Some research suggested that ADL disability had a more significant impact,^56^ while others argued the opposite.^57^ It was important to note that some studies might focus on only one of these variables, but at present, it remained unclear which level of functional disability had the most significant impact on depression. As we all knew, both ADL and IADL were indicators of physical functioning level in older adults, with IADL reflecting a higher level of independent living ability due to its inclusion of cognitive function assessment.^4,57^ To date, no studies had directly compared the effects of ADL and IADL disability on depression, which may suggest that such a comparison had limited practical significance. However, recognizing the bidirectional relationship between depression and functional disability, it was essential to pay attention to the potential mutual transformation of these two conditions. Therefore, comprehensive intervention strategies aimed at simultaneously improving physical function and mental health may be key to enhancing the quality of life for older adults.

Secondly, depression, as a type of mental disorder, was characterized by prolonged periods of low mood and was often accompanied by sleep disturbances ^63^. In severe cases, this could lead to sleep disorders, which in turn could exacerbate depressive symptoms, creating a vicious cycle. Existing research had confirmed the reciprocal relationship between depression and sleep,^27,28^ a connection that had been observed in cross-sectional and longitudinal studies.^28^ Most of the included studies had focused on exploring the impact of sleep as an exposure factor on depression. Evidences suggested that both excessive sleep and sleep deprivation could worsen depressive symptoms.^65^ However, most research indicated that short sleep duration or insomnia was a major risk factor for depression.^28,64^ Both long sleep and short sleep, as unhealthy sleep patterns, were considered as harmful to human health, affecting both gender equally. Consequently, addressing sleep problems may emerge as a crucial strategy for alleviating depression. Further research is needed to explore the specific mechanisms of this relationship and to develop effective treatment strategies.

Thirdly, current research on the relationship between sleep and functional disability was somewhat limited, which might be due to the fact that the connection between sleep and functional disability was influenced by a variety of factors, making it difficult to clearly establish the specific correlation between the two in the short term. However, existing studies had revealed that both excessive and insufficient sleep could have adverse effects on functional disability.^32^ It was notable that three studies had indicated a close connection between poor sleep quality and functional disability.^29,31,70^ Given that the physical functions of the elderly population continuously change with age, and the aging process is often accompanied by a range of health challenges, it was especially important to pay attention to and improve the sleep quality of older adults.

This review, incorporating existing research, examined the complex relationships and mechanisms of interaction among three key variables: depression, sleep and function disability. Depression might lead to sleep problems, which in turn might amplify depressive symptoms and further lead to functional disability. Conversely, functional disability might also affect sleep and depressive, creating a cycle of mutual influence. To achieve an in-depth comprehension of the dynamic relationships between these variables, we constructed a theoretical framework, highlighting sleep as a potential mediating factor in the bidirectional relationship between depression and functional disability. We proposed that sleep-related problems might not only be a consequence of depression and functional disability but also act as a catalyst for the interaction between the two. Through this theoretical framework, we aimed to provide a new perspective on understanding the complex interactions among these three variables and to offer a scientific basis for developing more effective intervention strategies, thereby improving the quality of life for individuals affected by depression, sleep problems, and functional disability.

The theoretical framework proposed by this study enhances our understanding of the complex relationship between depression, sleep, and functional disability, providing a new viewpoint on the interaction between mental health and physical health. By emphasizing the role of sleep as a potential mediating variable, this study offers a new explanatory pathway for understanding the bidirectional relationship between depression and functional disability. What’s more, the dynamic interaction network illustrates the mechanism of interaction among these three variables, providing a theoretical foundation and research direction for future studies.

The findings of this study can provide a more holistic perspective for clinicians, helping them to consider various factors when treating depression, sleep problems, and functional disability, and to develop more effective intervention strategies. By understanding the interaction among these three variables, more targeted intervention measures can be developed to improve the quality of life in older adults. Our review underscores the importance of interdisciplinary collaboration, prompting the joint effort of experts in psychology, psychiatry, sleep medicine, and rehabilitation medicine to address complex health issues.

Our review systematically examined the complex relationships between depression, sleep, and functional disability, highlighting the crucial role of these factors in the context of aging. Despite the comprehensive approach taken, there were several limitations to acknowledge. Firstly, our study design was primarily focused on quantitative studies, which was partly due to the limited availability of qualitative research, but also to some extent due to the additional exclusion criteria we introduced during the full-text screening phase. Secondly, there was a selection bias in the study samples, as most studies focused on specific populations or regions, which might not be generalized to a broader population. Moreover, some of the cohort studies included lacked sufficient follow-up time to observe long-term effects. Although included studies attempted to control for confounding factors, the possible remained that unmeasured or unobserved factors could influence the results. It was also significant that assessments of sleep, depression, and functional disability relied on subjective data, with a lack of objective measures, which could affect the accuracy of the findings. The theoretical framework we constructed considered the interactions between multiple variables, but the validation of these relationships mighty be limited by methodological and statistical analysis in practice.

## Conclusion

This review systematically reviewed the longitudinal bidirectional relationship between depression, sleep, and functional disability, and constructed a conceptual framework emphasizing the critical role of sleep in the complex interaction between mental and physical health in older adults. The research results of this paper have important guiding significance for clinical intervention, interdisciplinary collaboration, improving quality of life, and public health policy formulation. In the future, more high-quality research is needed to verify and improve this theoretical framework, and explore more effective intervention strategies to improve sleep quality and the quality of life in older adults.

## Acknowledgement

None.

## Contributors

HW and WG designed the study. HW, LW, and HC did the literature search. HW, LW, and HC did the literature screening and data extraction. HW did the data synthesis and wrote the manuscript. HW and LW revised the manuscript from preliminary draft to submission. WG supervised the study. All authors have read and approved the manuscript. WG is the guarantor.

## Patient and public involvement

Patients and/or the public were not involved in the design, conduct, reporting or dissemination plans of this research.

## Data availability statement

No data was used for the research described in the article.

