## Supplementary Appendix 2 for "The longitudinal mediating role of sleep in the bidirectional relationship between depression and functional disability among older adults: A systematic review and conceptual framework"

**Supplementary Table S1.** Databases search strategy.

| **Topic** | **Databases** | **Searches** | **Results** |
| --- | --- | --- | --- |
| **Dpression and functional disability** | **MEDLINE** | depression OR Patient Health Questionnaire OR Adjustment Disorders OR Affective Disorders, Psychotic OR Bipolar Disorder OR Depression OR Depressive Disorder, Major OR Depressive Disorder OR Seasonal Affective Disorder OR Dysthymic Disorder OR Depressive Disorder, Treatment-Resistant (MeSH Heading) AND Activities of Daily Living OR Disability Studies OR Developmental Disability Nursing OR Healthy Life Expectancy OR Disability Evaluation OR Disabled Persons OR International Classification of Functioning, Disability "AND" Health OR Mobility Limitation OR Independent Living OR Dependency, Psychological (MeSH Heading) | 8241 |
|  | **Pubmed** | ("Depression"[Mesh] OR "Depressive Disorder"[Mesh] OR "Depressive Disorder, Treatment-Resistant"[Mesh] OR "Bipolar Disorder"[Mesh] OR "Dysthymic Disorder"[Mesh] OR "Seasonal Affective Disorder"[Mesh] OR "Depressive Disorder, Major"[Mesh] OR "Adjustment Disorders"[Mesh] OR "Psychiatric Status Rating Scales"[Mesh] OR "Affective Disorders, Psychotic"[Mesh] OR "Major Depressive Disorder 1" [Supplementary Concept] OR "Major Depressive Disorder 2" [Supplementary Concept] OR "Patient Health Questionnaire"[Mesh]) AND ("International Classification of Functioning, Disability and Health"[Mesh]OR "Disability Studies"[Mesh] OR "Disabled Persons"[Mesh] OR "Physician Impairment"[Mesh] OR "Health Services for Persons with Disabilities"[Mesh] OR "Disability Evaluation"[Mesh] OR "Activities of Daily Living"[Mesh] OR "Mobility Limitation"[Mesh] OR "Dependency, Psychological" [Mesh] OR "Independent Living"[Mesh]) | 11535 |
|  | **Social Sciences Citation Index/ Science Citation Index/ ProQuest Dissertations and Theses Global** | (((((((TS=(depression)) OR TS=(affective disorders)) OR TS=(Bipolar depression)) OR TS=(major depression)) OR TS=(dysthymia)) OR TS=(adjustment disorders)) OR TS=(unipolar depression)) AND (((((((((((((TS=(functional disability)) OR TS=(functional limitation)) OR TS=(ADL)) OR TS=(IADL)) OR TS=(functional impairment)) OR TS=(physical disability)) OR TS=(mobility limitation)) OR TS=(activities of daily living)) OR TS=(dependency)) OR TS=(physical limitation)) OR TS=(physical impairment)) OR TS=(mobility disability)) OR TS=(activities disability)) OR TS=(instrumental activities of daily living) | 24595/ 34286/ 3162 |
|  | **Cochrane** | #1 MeSH descriptor: [Depression] explode all trees 18256 #2 MeSH descriptor: [Bipolar Disorder] explode all trees 3586 #3 MeSH descriptor: [Depressive Disorder, Major] explode all trees 7198 #4 MeSH descriptor: [Depressive Disorder] explode all trees 16626 #5 MeSH descriptor: [Depressive Disorder, Treatment-Resistant] explode all trees 754 #6 MeSH descriptor: [Mood Disorders] explode all trees 20282 #7 MeSH descriptor: [Affective Disorders, Psychotic] explode all trees 120 #8 MeSH descriptor: [Adjustment Disorders] explode all trees 285 #9 MeSH descriptor: [Affective Symptoms] explode all trees 584 #10 MeSH descriptor: [Dysthymic Disorder] explode all trees 200 #11 ("depression"):ti,ab,kw OR ("bipolar disorder"):ti,ab,kw OR ("depressive disorder, major"):ti,ab,kw OR ("depressive disorder"):ti,ab,kw OR ("depressive disorder, treatment-resistant"):ti,ab,kw 111022 #12 ("mood disorder"):ti,ab,kw OR ("affective disorders"):ti,ab,kw OR ("adjustment disorders"):ti,ab,kw OR ("affective symptoms"):ti,ab,kw OR ("dysthymic disorder"):ti,ab,kw 5081 #13 MeSH descriptor: [Disability Evaluation] explode all trees 4866 #14 MeSH descriptor: [Disabled Persons] explode all trees 1719 #15 MeSH descriptor: [Physician Impairment] explode all trees 8 #16 MeSH descriptor: [Activities of Daily Living] explode all trees 13239 #17 MeSH descriptor: [Mobility Limitation] explode all trees 690 #18 MeSH descriptor: [Dependency, Psychological] explode all trees 31 #19 MeSH descriptor: [Independent Living] explode all trees 1156 #20 MeSH descriptor: [Health Services for Persons with Disabilities] explode all trees 5 #21 MeSH descriptor: [Disability Studies] explode all trees 1 #22 ("disability evaluation"):ti,ab,kw OR ("disability"):ti,ab,kw OR ("disabled persons"):ti,ab,kw OR ("physician impairment"):ti,ab,kw OR ("activities of daily living"):ti,ab,kw 59745 #23 ("mobility limitation"):ti,ab,kw OR ("dependency"):ti,ab,kw OR ("independent living"):ti,ab,kw OR ("health services for persons with disabilities"):ti,ab,kw OR ("disability studies"):ti,ab,kw 6378 #24 #1 OR #2 OR #3 OR #4 OR #5 OR #6 OR #7 OR #8 OR #9 OR #10 OR #11 OR #12 113504 depression #25 #13 OR #14 OR #15 OR #16 OR #17 OR #18 OR #19 OR #20 OR #21 OR #22 OR #23 70634 functional disability #26 #24 AND #25 | 9522 |
|  | **Scopuse** | ( TITLE-ABS-KEY ( "depression" ) OR TITLE-ABS-KEY ( "major depressive disorder" ) OR TITLE-ABS-KEY ( "depressive symptoms" ) OR TITLE-ABS-KEY ( "depressive disorders" ) OR TITLE-ABS-KEY ( "bipolar disorder" ) OR TITLE-ABS-KEY ( "bipolar depression" ) OR TITLE-ABS-KEY ( "major depressive disorder" ) OR TITLE-ABS-KEY ( "major depression" ) OR TITLE-ABS-KEY ( "mood disorders" ) OR TITLE-ABS-KEY ( "affective disorders" ) OR TITLE-ABS-KEY ( "adjustment disorder" ) OR TITLE-ABS-KEY ( "affective symptoms" ) OR TITLE-ABS-KEY ( "dysthymic depression" ) OR TITLE-ABS-KEY ( "dysthymic disorder" ) ) AND ( TITLE-ABS-KEY ( "disability evaluation" ) OR TITLE-ABS-KEY ( "disabled persons" ) OR TITLE-ABS-KEY ( "functional disability" ) OR TITLE-ABS-KEY ( "functional limitation" ) OR TITLE-ABS-KEY ( "functional impairment" ) OR TITLE-ABS-KEY ( "mobility limitation" ) OR TITLE-ABS-KEY ( "disability" ) OR TITLE-ABS-KEY ( "disability studies" ) OR TITLE-ABS-KEY ( "physician impairment" ) OR TITLE-ABS-KEY ( "activities of daily living" ) OR TITLE-ABS-KEY ( "physical activity" ) OR TITLE-ABS-KEY ( "adl" ) OR TITLE-ABS-KEY ( "dependency" ) OR TITLE-ABS-KEY ( "independent living" ) ) | 68661 |
|  | **PsycArticles** | MeSH: Depression OR MeSH: depressive disorder OR MeSH: bipolar mood disorder OR MeSH: bipolar affective disorder OR MeSH: bipolar disorder OR MeSH: major depression OR MeSH: major depressive disorder OR MeSH: major affective disorder OR MeSH: mood disorder OR MeSH: affective disorder OR MeSH: adjustment disorder with depressed mood OR MeSH: affective symptoms OR MeSH: dysthymic disorder AND MeSH: functional disorder OR MeSH: functional limitation OR MeSH: functional independence measure OR MeSH: functional activities OR MeSH: functional impairment OR MeSH: disability OR MeSH: disability evaluation OR MeSH: disability syndrome OR MeSH: disabled personnel OR MeSH: physician impairment OR MeSH: physical activity OR MeSH: activities of daily living OR MeSH: mobility limitation OR MeSH: dependency OR MeSH: dependency need OR MeSH: independent living OR MeSH: health services for the aged | 127 |
| **Sleep and depression** | **Medline** | depression OR Patient Health Questionnaire OR Adjustment Disorders OR Affective Disorders, Psychotic OR Bipolar Disorder OR Depression OR Depressive Disorder, Major OR Depressive Disorder OR Seasonal Affective Disorder OR Dysthymic Disorder OR Depressive Disorder, Treatment-Resistant (MeSH Heading) AND Sleep Hygiene OR Sleep Latency OR Sleep, Slow-Wave OR Sleep Quality OR Sleep Initiation "AND" Maintenance Disorders OR Sleep OR Sleep Apnea Syndromes OR Sleep Deprivation OR Sleep Wake Disorders OR Sleep Stages OR Sleep, REM OR Polysomnography OR Sleep Disorders, Circadian Rhythm OR Sleep Apnea, Obstructive OR Sleep Apnea, Central OR Sleep Disorders, Intrinsic OR Dyssomnias OR Sleep Arousal Disorders OR Sleep-Wake Transition Disorders OR REM Sleep Parasomnias OR Lethargy (MeSH Heading) | 9543 |
|  | **Pubmed** | ("Sleep-Wake Transition Disorders"[Mesh] OR "Sleep Arousal Disorders"[Mesh] OR "Sleep Disorders, Intrinsic"[Mesh] OR "REM Sleep Behavior Disorder"[Mesh] OR "Sleep Disorders, Circadian Rhythm"[Mesh] OR "Sleep, REM"[Mesh] OR "Sleep Wake Disorders"[Mesh] OR "Sleep Deprivation"[Mesh] OR "Sleep Initiation and Maintenance Disorders"[Mesh] OR "Sleep Duration"[Mesh] OR "Sleep Quality"[Mesh] OR "Sleep Latency"[Mesh] OR "Sleep Hygiene"[Mesh] OR "Dyssomnias"[Mesh] OR "Parasomnias"[Mesh] OR "Sleep"[Mesh]) AND ("Depression"[Mesh] OR "Depressive Disorder"[Mesh] OR "Depressive Disorder, Treatment-Resistant"[Mesh] OR "Bipolar Disorder"[Mesh] OR "Dysthymic Disorder"[Mesh] OR "Seasonal Affective Disorder"[Mesh] OR "Depressive Disorder, Major"[Mesh] OR "Adjustment Disorders"[Mesh] OR "Psychiatric Status Rating Scales"[Mesh] OR "Affective Disorders, Psychotic"[Mesh] OR "Major Depressive Disorder 1" [Supplementary Concept] OR "Major Depressive Disorder 2" [Supplementary Concept] OR "Patient Health Questionnaire"[Mesh]) | 10191 |
|  | **Social Sciences Citation Index/ Science Citation Index/ ProQuest Dissertations and Theses Global** | ((TS=(sleep)) OR TS=(REM)) AND ((((((TS=(depression)) OR TS=(affective disorders)) OR TS=(Bipolar depression)) OR TS=(major depression)) OR TS=(dysthymia)) OR TS=(adjustment disorders)) OR TS=(unipolar depression) | 22214/ 37437/ 1855 |
|  | **Cochrane** | #27 MeSH descriptor: [Sleep] explode all trees 8810 #28 MeSH descriptor: [Sleep Apnea Syndromes] explode all trees 4129 #29 MeSH descriptor: [Sleep Wake Disorders] explode all trees 12387 #30 MeSH descriptor: [Dyssomnias] explode all trees 10259 #31 MeSH descriptor: [Sleep Arousal Disorders] explode all trees 20 #32 MeSH descriptor: [Sleep Disorders, Circadian Rhythm] explode all trees 271 #33 MeSH descriptor: [Sleep Disorders, Intrinsic] explode all trees 8699 #34 MeSH descriptor: [Sleep Deprivation] explode all trees 1115 #35 MeSH descriptor: [Sleep Latency] explode all trees 31 #36 MeSH descriptor: [REM Sleep Behavior Disorder] explode all trees 48 #37 MeSH descriptor: [Sleep, REM] explode all trees 738 #38 MeSH descriptor: [Sleep Initiation and Maintenance Disorders] explode all trees 3751 #39 MeSH descriptor: [Sleep Duration] explode all trees 21 #40 MeSH descriptor: [Sleep Quality] explode all trees 325 #41 MeSH descriptor: [Sleep Hygiene] explode all trees 399 #42 ("sleep-apnea syndromes"):ti,ab,kw OR ("sleep"):ti,ab,kw OR ("sleep wake disorders"):ti,ab,kw OR ("dyssomnias"):ti,ab,kw OR ("sleep arousal disorders"):ti,ab,kw 53901 #43 ("sleep disorders,circadian rhythm"):ti,ab,kw OR ("sleep disorders,intrinsic"):ti,ab,kw OR ("sleep deprivation"):ti,ab,kw OR ("sleep latency"):ti,ab,kw OR ("sleep,REM"):ti,ab,kw 5175 #44 ("REM sleep behavior disorder"):ti,ab,kw OR ("sleep initiation and maintenance disorders"):ti,ab,kw OR ("sleep duration"):ti,ab,kw OR ("sleep quality"):ti,ab,kw OR ("sleep hygiene"):ti,ab,kw 18989 #45 #27 OR #28 OR #29 OR #30 OR #31 OR #32 OR #33 OR #34 OR #35 OR #36 OR #37 OR #38 OR #39 OR #40 OR #41 OR #42 OR #43 OR #44 54469 sleep #46 #45 AND #24 | 11897 |
|  | **Scopuse** | ( TITLE-ABS-KEY ( "sleep" ) OR TITLE-ABS-KEY ( "sleep disorders" ) OR TITLE-ABS-KEY ( "sleep apnea" ) OR TITLE-ABS-KEY ( "sleep deprivation" ) OR TITLE-ABS-KEY ( "sleep latency" ) OR TITLE-ABS-KEY ( "sleep duration" ) OR TITLE-ABS-KEY ( "sleep quality" ) OR TITLE-ABS-KEY ( "REM sleep" ) OR TITLE-ABS-KEY ( "REM sleep behavior disorder" ) OR TITLE-ABS-KEY ( "sleep hygiene" ) OR TITLE-ABS-KEY ( "sleep initiation and maintenance disorders" ) OR TITLE-ABS-KEY ( "dyssomnia" ) OR TITLE-ABS-KEY ( "sleep disturbance" ) ) AND ( TITLE-ABS-KEY ( "depression" ) OR TITLE-ABS-KEY ( "major depressive disorder" ) OR TITLE-ABS-KEY ( "depressive symptoms" ) OR TITLE-ABS-KEY ( "depressive disorders" ) OR TITLE-ABS-KEY ( "bipolar disorder" ) OR TITLE-ABS-KEY ( "bipolar depression" ) OR TITLE-ABS-KEY ( "major depressive disorder" ) OR TITLE-ABS-KEY ( "major depression" ) OR TITLE-ABS-KEY ( "mood disorders" ) OR TITLE-ABS-KEY ( "affective disorders" ) OR TITLE-ABS-KEY ( "adjustment disorder" ) OR TITLE-ABS-KEY ( "affective symptoms" ) OR TITLE-ABS-KEY ( "dysthymic depression" ) OR TITLE-ABS-KEY ( "dysthymic disorder" ) ) | 58512 |
|  | **PsycArticles** | MeSH: Depression OR MeSH: depressive disorder OR MeSH: bipolar mood disorder OR MeSH: bipolar affective disorder OR MeSH: bipolar disorder OR MeSH: major depression OR MeSH: major depressive disorder OR MeSH: major affective disorder OR MeSH: mood disorder OR MeSH: affective disorder OR MeSH: adjustment disorder with Depression OR MeSH: depressive disorder OR MeSH: bipolar mood disorder OR MeSH: bipolar affective disorder OR MeSH: bipolar disorder OR MeSH: major depression OR MeSH: major depressive disorder OR MeSH: major affective disorder OR MeSH: mood disorder OR MeSH: affective disorder OR MeSH: adjustment disorder with depressed mood OR MeSH: affective symptoms OR MeSH: dysthymic disorder AND MeSH: Sleep OR MeSH: sleep arousal disorders OR MeSH: sleep deprivation OR MeSH: sleep disorder OR MeSH: sleep apnea OR MeSH: sleep apnea syndromes OR MeSH: sleep latency OR MeSH: REM sleep OR MeSH: REM sleep behavior disorder OR MeSH: sleep hygiene OR MeSH: sleep duration OR MeSH: sleep quality OR MeSH: “sleep initiation and maintenance disorders” | 148 |
| **Sleep and functional disability** | **Medline** | Sleep Hygiene OR Sleep Latency OR Sleep, Slow-Wave OR Sleep Quality OR Sleep Initiation "AND" Maintenance Disorders OR Sleep OR Sleep Apnea Syndromes OR Sleep Deprivation OR Sleep Wake Disorders OR Sleep Stages OR Sleep, REM OR Polysomnography OR Sleep Disorders, Circadian Rhythm OR Sleep Apnea, Obstructive OR Sleep Apnea, Central OR Sleep Disorders, Intrinsic OR Dyssomnias OR Sleep Arousal Disorders OR Sleep-Wake Transition Disorders OR REM Sleep Parasomnias OR Lethargy (MeSH Heading) AND Activities of Daily Living OR Disability Studies OR Developmental Disability Nursing OR Healthy Life Expectancy OR Disability Evaluation OR Disabled Persons OR International Classification of Functioning, Disability "AND" Health OR Mobility Limitation OR Independent Living OR Dependency, Psychological (MeSH Heading) | 1896 |
|  | **Pubmed** | ("Sleep-Wake Transition Disorders"[Mesh] OR "Sleep Arousal Disorders"[Mesh] OR "Sleep Disorders, Intrinsic"[Mesh] OR "REM Sleep Behavior Disorder"[Mesh] OR "Sleep Disorders, Circadian Rhythm"[Mesh] OR "Sleep, REM"[Mesh] OR "Sleep Wake Disorders"[Mesh] OR "Sleep Deprivation"[Mesh] OR "Sleep Initiation and Maintenance Disorders"[Mesh] OR "Sleep Duration"[Mesh] OR "Sleep Quality"[Mesh] OR "Sleep Latency"[Mesh] OR "Sleep Hygiene"[Mesh] OR "Dyssomnias"[Mesh] OR "Parasomnias"[Mesh] OR "Sleep"[Mesh]) AND ("International Classification of Functioning, Disability and Health"[Mesh]OR "Disability Studies"[Mesh] OR "Disabled Persons"[Mesh] OR "Physician Impairment"[Mesh] OR "Health Services for Persons with Disabilities"[Mesh] OR "Disability Evaluation"[Mesh] OR "Activities of Daily Living"[Mesh] OR "Mobility Limitation"[Mesh] OR "Dependency, Psychological" [Mesh] OR "Independent Living"[Mesh]) | 2114 |
|  | **Social Sciences Citation Index/ Science Citation Index/ ProQuest Dissertations and Theses Global** | ((TS=(sleep)) OR TS=(REM)) AND (((((((((((((TS=(functional disability)) OR TS=(functional limitation)) OR TS=(ADL)) OR TS=(IADL)) OR TS=(functional impairment)) OR TS=(physical disability)) OR TS=(mobility limitation)) OR TS=(activities of daily living)) OR TS=(dependency)) OR TS=(physical limitation)) OR TS=(physical impairment)) OR TS=(mobility disability)) OR TS=(activities disability)) OR TS=(instrumental activities of daily living) | 8220/ 14005/ 1150 |
|  | **Cochrane** | #47 #45 AND #25 | 2683 |
|  | **Scopuse** | ( TITLE-ABS-KEY ( "sleep" ) OR TITLE-ABS-KEY ( "sleep disorders" ) OR TITLE-ABS-KEY ( "sleep apnea" ) OR TITLE-ABS-KEY ( "sleep deprivation" ) OR TITLE-ABS-KEY ( "sleep latency" ) OR TITLE-ABS-KEY ( "sleep duration" ) OR TITLE-ABS-KEY ( "sleep quality" ) OR TITLE-ABS-KEY ( "REM sleep" ) OR TITLE-ABS-KEY ( "REM sleep behavior disorder" ) OR TITLE-ABS-KEY ( "sleep hygiene" ) OR TITLE-ABS-KEY ( "sleep initiation and maintenance disorders" ) OR TITLE-ABS-KEY ( "dyssomnia" ) OR TITLE-ABS-KEY ( "sleep disturbance" ) ) AND ( TITLE-ABS-KEY ( "disability evaluation" ) OR TITLE-ABS-KEY ( "disabled persons" ) OR TITLE-ABS-KEY ( "functional disability" ) OR TITLE-ABS-KEY ( "functional limitation" ) OR TITLE-ABS-KEY ( "functional impairment" ) OR TITLE-ABS-KEY ( "mobility limitation" ) OR TITLE-ABS-KEY ( "disability" ) OR TITLE-ABS-KEY ( "disability studies" ) OR TITLE-ABS-KEY ( "physician impairment" ) OR TITLE-ABS-KEY ( "activities of daily living" ) OR TITLE-ABS-KEY ( "physical activity" ) OR TITLE-ABS-KEY ( "adl" ) OR TITLE-ABS-KEY ( "dependency" ) OR TITLE-ABS-KEY ( "independent living" ) ) | 29822 |
|  | **PsycArticles** | MeSH: functional disorder OR MeSH: functional limitation OR MeSH: functional independence measure OR MeSH: functional activities OR MeSH: functional impairment OR MeSH: disability OR MeSH: disability evaluation OR MeSH: disability syndrome OR MeSH: disabled personnel OR MeSH: physician impairment OR MeSH: physical activity OR MeSH: activities of daily living OR MeSH: mobility limitation OR MeSH: dependency OR MeSH: dependency need OR MeSH: independent living OR MeSH: health services for the aged AND MeSH: Sleep OR MeSH: sleep arousal disorders OR MeSH: sleep deprivation OR MeSH: sleep disorder OR MeSH: sleep apnea OR MeSH: sleep apnea syndromes OR MeSH: sleep latency OR MeSH: REM sleep OR MeSH: REM sleep behavior disorder OR MeSH: sleep hygiene OR MeSH: sleep duration OR MeSH: sleep quality OR MeSH: “sleep initiation and maintenance disorders” | 108 |
| **Sleep, depression, and functional disability** | **Medline** | depression OR Patient Health Questionnaire OR Adjustment Disorders OR Affective Disorders, Psychotic OR Bipolar Disorder OR Depression OR Depressive Disorder, Major OR Depressive Disorder OR Seasonal Affective Disorder OR Dysthymic Disorder OR Depressive Disorder, Treatment-Resistant (MeSH Heading) AND Sleep Hygiene OR Sleep Latency OR Sleep, Slow-Wave OR Sleep Quality OR Sleep Initiation "AND" Maintenance Disorders OR Sleep OR Sleep Apnea Syndromes OR Sleep Deprivation OR Sleep Wake Disorders OR Sleep Stages OR Sleep, REM OR Polysomnography OR Sleep Disorders, Circadian Rhythm OR Sleep Apnea, Obstructive OR Sleep Apnea, Central OR Sleep Disorders, Intrinsic OR Dyssomnias OR Sleep Arousal Disorders OR Sleep-Wake Transition Disorders OR REM Sleep Parasomnias OR Lethargy (MeSH Heading) AND Activities of Daily Living OR Disability Studies OR Developmental Disability Nursing OR Healthy Life Expectancy OR Disability Evaluation OR Disabled Persons OR International Classification of Functioning, Disability "AND" Health OR Mobility Limitation OR Independent Living OR Dependency, Psychological (MeSH Heading) | 371 |
|  | **Pubmed** | ("Sleep-Wake Transition Disorders"[Mesh] OR "Sleep Arousal Disorders"[Mesh] OR "Sleep Disorders, Intrinsic"[Mesh] OR "REM Sleep Behavior Disorder"[Mesh] OR "Sleep Disorders, Circadian Rhythm"[Mesh] OR "Sleep, REM"[Mesh] OR "Sleep Wake Disorders"[Mesh] OR "Sleep Deprivation"[Mesh] OR "Sleep Initiation and Maintenance Disorders"[Mesh] OR "Sleep Duration"[Mesh] OR "Sleep Quality"[Mesh] OR "Sleep Latency"[Mesh] OR "Sleep Hygiene"[Mesh] OR "Dyssomnias"[Mesh] OR "Parasomnias"[Mesh] OR "Sleep"[Mesh]) AND ("International Classification of Functioning, Disability and Health"[Mesh]OR "Disability Studies"[Mesh] OR "Disabled Persons"[Mesh] OR "Physician Impairment"[Mesh] OR "Health Services for Persons with Disabilities"[Mesh] OR "Disability Evaluation"[Mesh] OR "Activities of Daily Living"[Mesh] OR "Mobility Limitation"[Mesh] OR "Dependency, Psychological" [Mesh] OR "Independent Living"[Mesh]) AND ("Depression"[Mesh] OR "Depressive Disorder"[Mesh] OR "Depressive Disorder, Treatment-Resistant"[Mesh] OR "Bipolar Disorder"[Mesh] OR "Dysthymic Disorder"[Mesh] OR "Seasonal Affective Disorder"[Mesh] OR "Depressive Disorder, Major"[Mesh] OR "Adjustment Disorders"[Mesh] OR "Psychiatric Status Rating Scales"[Mesh] OR "Affective Disorders, Psychotic"[Mesh] OR "Major Depressive Disorder 1" [Supplementary Concept] OR "Major Depressive Disorder 2" [Supplementary Concept] OR "Patient Health Questionnaire"[Mesh]) | 447 |
|  | **Social Sciences Citation Index/ Science Citation Index/ ProQuest Dissertations and Theses Global** | ((TS=(sleep)) OR TS=(REM)) AND (((((((TS=(depression)) OR TS=(affective disorders)) OR TS=(Bipolar depression)) OR TS=(major depression)) OR TS=(dysthymia)) OR TS=(adjustment disorders)) OR TS=(unipolar depression)) AND (((((((((((((TS=(functional disability)) OR TS=(functional limitation)) OR TS=(ADL)) OR TS=(IADL)) OR TS=(functional impairment)) OR TS=(physical disability)) OR TS=(mobility limitation)) OR TS=(activities of daily living)) OR TS=(dependency)) OR TS=(physical limitation)) OR TS=(physical impairment)) OR TS=(mobility disability)) OR TS=(activities disability)) OR TS=(instrumental activities of daily living) | 5876/ 8272/ 562 |
|  | **Cochrane** | #48 #24 AND #25 AND #45 | 1047 |
|  | **Scopuse** | ( TITLE-ABS-KEY ( "sleep" ) OR TITLE-ABS-KEY ( "sleep disorders" ) OR TITLE-ABS-KEY ( "sleep apnea" ) OR TITLE-ABS-KEY ( "sleep deprivation" ) OR TITLE-ABS-KEY ( "sleep latency" ) OR TITLE-ABS-KEY ( "sleep duration" ) OR TITLE-ABS-KEY ( "sleep quality" ) OR TITLE-ABS-KEY ( "REM sleep" ) OR TITLE-ABS-KEY ( "REM sleep behavior disorder" ) OR TITLE-ABS-KEY ( "sleep hygiene" ) OR TITLE-ABS-KEY ( "sleep initiation and maintenance disorders" ) OR TITLE-ABS-KEY ( "dyssomnia" ) OR TITLE-ABS-KEY ( "sleep disturbance" ) ) AND ( TITLE-ABS-KEY ( "depression" ) OR TITLE-ABS-KEY ( "major depressive disorder" ) OR TITLE-ABS-KEY ( "depressive symptoms" ) OR TITLE-ABS-KEY ( "depressive disorders" ) OR TITLE-ABS-KEY ( "bipolar disorder" ) OR TITLE-ABS-KEY ( "bipolar depression" ) OR TITLE-ABS-KEY ( "major depressive disorder" ) OR TITLE-ABS-KEY ( "major depression" ) OR TITLE-ABS-KEY ( "mood disorders" ) OR TITLE-ABS-KEY ( "affective disorders" ) OR TITLE-ABS-KEY ( "adjustment disorder" ) OR TITLE-ABS-KEY ( "affective symptoms" ) OR TITLE-ABS-KEY ( "dysthymic depression" ) OR TITLE-ABS-KEY ( "dysthymic disorder" ) ) AND ( TITLE-ABS-KEY ( "disability evaluation" ) OR TITLE-ABS-KEY ( "disabled persons" ) OR TITLE-ABS-KEY ( "functional disability" ) OR TITLE-ABS-KEY ( "functional limitation" ) OR TITLE-ABS-KEY ( "functional impairment" ) OR TITLE-ABS-KEY ( "mobility limitation" ) OR TITLE-ABS-KEY ( "disability" ) OR TITLE-ABS-KEY ( "disability studies" ) OR TITLE-ABS-KEY ( "physician impairment" ) OR TITLE-ABS-KEY ( "activities of daily living" ) OR TITLE-ABS-KEY ( "physical activity" ) OR TITLE-ABS-KEY ( "adl" ) OR TITLE-ABS-KEY ( "dependency" ) OR TITLE-ABS-KEY ( "independent living" ) ) | 8720 |
|  | **PsycArticles** | MeSH: Depression OR MeSH: depressive disorder OR MeSH: bipolar mood disorder OR MeSH: bipolar affective disorder OR MeSH: bipolar disorder OR MeSH: major depression OR MeSH: major depressive disorder OR MeSH: major affective disorder OR MeSH: mood disorder OR MeSH: affective disorder OR MeSH: adjustment disorder with Depression OR MeSH: depressive disorder OR MeSH: bipolar mood disorder OR MeSH: bipolar affective disorder OR MeSH: bipolar disorder OR MeSH: major depression OR MeSH: major depressive disorder OR MeSH: major affective disorder OR MeSH: mood disorder OR MeSH: affective disorder OR MeSH: adjustment disorder with depressed mood OR MeSH: affective symptoms OR MeSH: dysthymic disorder AND MeSH: Sleep OR MeSH: sleep arousal disorders OR MeSH: sleep deprivation OR MeSH: sleep disorder OR MeSH: sleep apnea OR MeSH : sleep apnea syndromes OR MeSH: sleep latency OR MeSH: REM sleep OR MeSH: REM sleep behavior disorder OR MeSH: sleep hygiene OR MeSH: sleep duration OR MeSH: sleep quality OR MeSH: “sleep initiation and maintenance disorders” AND MeSH: functional disorder OR MeSH: functional limitation OR MeSH: functional independence measure OR MeSH: functional activities OR MeSH: functional impairment OR MeSH: disability OR MeSH: disability evaluation OR MeSH: disability syndrome OR MeSH: disabled personnel OR MeSH: physician impairment OR MeSH: physical activity OR MeSH: activities of daily living OR MeSH: mobility limitation OR MeSH: dependency OR MeSH: dependency need OR MeSH: independent living OR MeSH: health services for the aged | 70 |
|  |  |  | 397289 |

**Supplementary Table S2.** Characteristics of included studies.

| **References** | **Publication date** | **Study design** | **Aim/research question** | **Data collection** | **Sample size** | **Exposure measures** | **Outcome measures** | **Follow-up time** | **Effect sizes** | **Summary of main findings** |
| --- | --- | --- | --- | --- | --- | --- | --- | --- | --- | --- |
| **The relationship between depression and functional disability** | | | | | | | | | | |
| Li Wang et al., 2002 | 2002/1/1 | Cohort study. | To identify factors associated with functional change in an older population and investigate interactions among selected potential risk factors. | Data are from the Adult Changes in Thought (ACT) Study. | 2,581 remained in the study. | Chronic medical conditions and other nonmedical factors. | Functional status. | 4 years. | Over 4 years of follow-up,depression were associated with increased age-adjusted rate of functional decline in ADLs [0.08 (0.01–0.15)], IADLs [0.09 (0.01–0.16)], and PPF [-0.30 (-0.41 to -0.19)]. | Over the follow-up period, coronary heart disease, CVD, and depression were associated with increased rates of functional decline. |
| Milan Chang et al., 2009 | 2009/4/1 | Cohort study. | To examine whether onset of worsening disability has an impact on depressive symptoms over a short time interval among older women with mild to severe disability. | Data from the Women’s Health and Aging Study (WHAS). | A total of 671 subjects were eligible for the study. | ADLs disability. | Depressive symptoms. | 3 years. | In a model that used all rounds of data and adjusted for multiple confounders, women with worsening disability were more likely to develop depressive symptoms at the time of worsening disability compared with those who remained stable (OR 2.2, 95%CI 1.1-4.3). For those developing new disability without depressive symptoms, the adjusted OR for developing depressive symptoms 6 months later was 1.7 (95%CI 0.6-4.8). | The impact of worsening disability on depressive symptoms among older women with mild to severe disability living in the community was examined at the same time as subjects reported worsening disability, and then 6 months later. |
| Kenneth E. Covinsky et al., 2010 | 2010/3/1 | Cohort study. | The study examined whether subjects with high levels of depressive symptoms were more likely to develop persistent difficulty in ADLs and mobility functioning over 12 years as these subjects entered old age. | A study was conducted in the Health and Retirement Study (HRS). | A final sample size of 7,207 subjects. | Depressive symptoms. | Basic mobility and ADL tasks. | 12 years. | Over 12 years of follow-up, subjects with depressive symptoms were more likely to reach the primary outcome measure of persistent difficulty with mobility or ADL function (Cox HR=1.44, 95%CI=1.25–1.66). | It was found that middle-aged subjects with depressive symptoms were more likely than those without to develop difficulty with mobility and ADLs over 12 years of follow-up. |
| Celia F. Hybels et al.,2009 | 2009/10/1 | Cohort study. | To to determine the importance of subthreshold depression by exploring the incremental effect of more symptomatic depression over the effects of lower levels of depressive symptomatology predicting functional change and to model the functional form of the relationship between depressive symptoms and functional change across the three outcomes. | The data derive from the Duke Established Populations for Epidemiologic Studies of the Elderly (EPESE). | An analysis sample of 6432 observations for the mixed models. | Depressive symptoms. | Basic ADL tasks, IADL tasks and mobility. | 10 years. | Race, sex, age, education, marital status, cognitive status, health status, self-perceived health, perceived social support, and functional status at the index interview, having 6+ depressive symptoms predicted an increase of 0.12 IADL limitations 3–4 years later (p=0.03). | We conclude the effect of depression on decline in IADL abilities occurs at low levels of depressive symptomatology, and that more symptoms may not confer significantly greater risk of decline. And we conclude the relationship between depressive symptoms and functional change is complex, may differ by domain of function assessed, and may not necessarily be linear. |
| Hajime Iwasa et al., 2009 | 2009/11/1 | Cohort study. | This study aimed to examine a longitudinal relationship between depression status and functional decline among Japanese community-dwelling older adults, using a 12-year population-based, prospective cohort study design. | The source of data for the present study was the Longitudinal Interdisciplinary Study on Aging conducted by the Tokyo Metropolitan Institute of Gerontology. | In total, 710 participants (283 men and 427 women) with a complete data set were included, and their data were used for the 12-year follow-up. | Depression status. | Functional capacity. | 12 years. | Following multivariate Cox regression analysis, adjusted for the potential confounders cited above, depression status was significantly and independently associated with BADL decline (RR=1.46, 95%CI:1.13–1.89) and with higher-level competence decline (RR=1.56, 95%CI: 1.18–2.04). | Depression status predicted BADL decline and higher-level competence decline, and also identified linear relationships between the level of depression status and functional decline (both BADL decline and higher-level competence decline). |
| Carolyn L. Turvey et al., 2009 | 2009/8/1 | Cohort study. | This study examines the relative contribution of cognitive function, physical function, and chronic illness to depression two years later in a nationwide sample of elders aged 70 and older. | Data from the Asset and Health Dynamics Among the Oldest Old (AHEAD). | A total of 5289 elders completing two waves in this study. | Cognitive function; high blood pressure, diabetes, lung disease, heart disease, arthritis; activities of daily living (ADL’s) and instrumental activities of daily living (IADL’s). | Depression symptoms. | 2 years. | In a full multivariate model, baseline ADLs and IADLs predicted depression as measured by the CESD-8 at Wave 2 (ADLs: RR=0.10, P<0.01; IADLs: RR=0.05, P=0.03). | Chronic illness, physical function, and cognitive function all independently predict depressive morbidity in late-life. |
| Mark I. Weinberger et al., 2009 | 2009/9/1 | Cohort study. | Our primary aim was to identify baseline clinical and functional factors associated with the risk of depression at one-year follow-up. | Data for the current study were collected as part of a longitudinal study on the prevalence, course, and outcomes of major depression in elderly patients receiving home-care skilled nursing services. | We included 268 patients interviewed at one year. | Disabilities in activities of daily living (ADLs), instrumental activities of daily living (IADLs), and mobility. | Depression symptoms. | 1 year. | In multivariate analyses, we found that worse self-rated health (OR=0.53, p=0.042), more somatic depressive symptoms (OR=1.19, p=0.015), greater number of ADL limitations at baseline (OR=1.63, p=0.014) and greater decline in ADL functioning from baseline to one year (OR=1.59, p=0.022) were all independently associated with onset depression. | Older disabled adults with poor self-rated health, mild somatic depressive symptoms, a greater number of ADL limitations at baseline and declining ADL functioning are at risk of developing a depressive episode. |
| Mathew D. Gayman et al., 2008 | 2008/7/1 | Cohort study. | To assess the independent significance of prior level of physical limitations in predicting changes in depressive symptoms, and vice versa, considering the mediating effects of intervening measures of pain and stress exposure. | The data employed in this study were from a two-wave (W1 and W2) panel study of Miami-Dade County residents that included a substantial oversampling of individuals with a physical disability. | A total of 1,455 study participants provided valid data across all study variables employed in the current analysis. | Physical limitations. | Depressive symptomatology. | 3 years. | Although the level of physical limitations at W1 predicted changes in depressive symptoms (b=0.11, p< 0.001), these results offered no evidence that prior depressive symptoms were of significance in predicting changes in physical limitations. | We concluded that most of whatever causation may have been involved in the clear relationship between physical limitations and depressive symptomatology over a 3-year period flowed from physical limitations to depressive symptoms rather than in the reverse direction. We proposed that the association between physical limitations and depressive symptoms would be mediated by our indicators of pain and social stress measured at W2. |
| Naoki Kondo et al., 2008 | 2008/5/1 | Cohort study. | We investigated the impact of mental health on the decline in higher activities of daily living (ADL) defined in terms of social role performance (SR, the highest ADL), intellectual activity (IA), and instrumental ADL (IADL), as well as the onset of basic ADL disability. | The baseline data were collected from a two-stage probability sample of 1800 non-institutionalized older adults (65 or older) in Yamanashi prefecture in 2002. | 581 samples were used for analysis. | Mental health. | Higher ADL decline and onset of BADL disability. | 25 months. | Among the young–old subjects, severe depressive symptoms strongly predicted subsequent higher IADL decline: the adjusted HR (95% CI) was 4.06 (1.23,13.3), whereas the adjusted HR for SR and IA decline was 1.03 (0.12, 8.63) and 0.97 (0.24, 3.98). Severe depressive symptoms were strongly associated with subsequent higher ADL decline in the old–old subject, those with severe depressive symptoms showed the adjusted hazard ratios (HR) (95% confidence intervals: CI) of 3.22 (1.35, 7.71), 3.11 (1.38, 6.98), and 2.41 (1.07, 5.40) for the decline in SR, IA, and IADL, respectively. | The present study provides important evidence that severely depressive older persons have two- to three-fold higher risk for higher ADL decline compared to nondepressive persons and this excess risk was partly explained by social inactivity. |
| Jason E. Schillerstro et al., 2008 | 2008/9/1 | Review. | The goal of this article is to describe the intermediate pathways that may link depression with disability. | Published studies were identified through searches of the MEDLINE and PsychINFO databases from periods March 1966 to 2006. | Of the 20 studies reviewed. | Depression. | Functional status. |  | For positive studies, adjusted odds ratios measuring the longitudinal association between depression and disability ranged from 1.16 (95%CI 1.13-1.19) to 5.47 (95%CI 1.77-16.92). | From these longitudinal studies we conclude that both baseline depression and incident depression are independent predictors of functional decline in elders regardless of how either is assessed. |
| YANG YANG, 2016 | 2006/12/1 | Cohort study. | This study examines the process whereby functional disability amplifies depressive symptoms through decreasing perceived social support and psychological resources. | Longitudinal data for this study are from the North Carolina Established Populations for Epidemiologic Studies of the Elderly (EPESE). | This leaves an effective sample size of 1,149. | Functional disability status, social support, sense of control, and self-esteem. | Depressive symptoms. | 6 years. | The diagonal line linking Disability (T1) and CES-D (T2) indicates the significant direct effect of baseline disability (0.09, p < .001) on increases in CES-D from T1 to T2. The total effect of Disability (T1) on changes in CES-D is 0.22, 8 of which 40.7 percent is its direct effect(0.09/0.22). | The results of longitudinal change models and path analyses show that the perceived availability of a confidant, satisfaction with support, sense of control, and self-esteem mediate the effects of disability on increments in depressive symptoms in late life. |
| Dorothy D. Dunlop et al., 2005 | 2005/11/1 | Cohort study. | We evaluated the effect of depression on risk, on the basis of standardized assessment, for developing activities of daily living (ADL) disability. | Data from the Health and Retirement Study (HRS). | We limited our analyses to a cohort of 6871 older adults without baseline ADL task limitations who lived for at least 2 years after the initial interview. | Depression. | ADL disability. | 2 years. | Depression remained a strong and significant risk factor (adjusted odds ratio = 2.0; CI = 1.3, 3.1) for the development of ADL disability. | We have provided evidence of a substantial national public health burden related to depression in the development of ADL disability among US adults aged 54–65 years. Nearly 1 of every 5 new cases of ADL disability is associated with depression. |
| Nicholas P. Emptage et al., 2005 | 2005/4/1 | Cohort study. | We analyzed whether individuals with depression and comorbid pain in the Health and Retirement Study were more likely than those with depression alone to become physically disabled. | Data from the Health and Retirement Study (HRS), a longitudinal national survey of individuals born between 1931 and 1941. | Data were available for 8,280 respondents at baseline. | Depression, pain. | Imitations in activities of daily living. | 6 years. | Compared with the group with depression alone, the group with depression plus mild or moderate pain had a significantly greater probability of a new limitation for all the follow-up years (1996, χ2 =34.21, df=1, p<0.01; 1998, χ2= 38.26, df=1, p<0.01;2000, χ2 =25.22, df=1, p<0.01). The probability of a new limitation was also significantly greater in the group with depression plus severe pain compared with the group with depression alone for all the follow-up years (1996, χ2=29.25, df=1, p<0.01;1998, χ2=33.04, df=1, p<0.01; 2000,χ2=32.10, df=1, p<0.01). | After the analyses controlled for individual sociodemographic characteristics and medical conditions at baseline, compared with depression alone, depression plus pain—either mild or moderate pain or severe pain—was associated with significantly higher rates of new limitations in activities of daily living and work-limiting health problems over time. |
| Kala M. Mehta et al., 2002 | 2002/6/1 | Cohort study. | The purpose of this study is to determine the relative contributions of cognitive impairment and depressive symptoms on decline in activity of daily living (ADL) function over 2 years in an older cohort. | A U.S. national prospective cohort study of older people, Asset and Health Dynamics in the Oldest Old. | Five thousand six hundred ninety-seven participants (mean age 77, 64% women, 86% white) followed from 1993 to 1995. | Depressive symptoms and cognitive impairment. | Functional decline. | 2 years. | Depressed individuals were twice as likely to have subsequent decline as nondepressed individuals (RR=2.1, 95%CI=1.7–2.6).For participants who were independent at baseline, cognitive impairment (RR=2.3, 95%CI=1.7–3.1) and depressive symptoms (RR=1.9, 95%CI=1.3–2.6) together predicted the risk of incident functional decline. | Cognitive impairment and depressive symptoms are independent predictors of ADL dependence in participants who are independent in all ADLs at baseline. Cognitive impairment and depressive symptoms were robust risk factors; those with both risk factors had a slightly greater risk of decline than those with either risk factor alone. |
| Jason Schnittker, 2005 | 2005/1/1 | Cohort study. | The paper examines the effects of chronic illness and disability on depressive symptoms and examines whether the relationship between illness and disability and depressive symptoms changes with age. | Data for this study are from two longitudinal studies, the Health and Retirement Study (HRS) and the Study of Assets and Health Dynamics Among the Oldest Old (AHEAD). | 22,293 were included in at least one wave of the sample. | Illness, and functional disability. | Depression symptoms. | 8-9 years. | Activities in daily living (*b*=3.006, P<0.01), mobility (b=1.626, P<0.01, and strength (b=1.863, P<0.01) increased depressive symptoms. | Three forms of disability (activities in daily living, mobility, and strength) substantially increase depressive symptoms. Disability leads to more depressive symptoms when experienced at younger ages. |
| YANG YANG et al., 2005 | 2005/6/1 | Cohort study. | This article addresses how stable functional disability statuses and disability transitions are related to change in depressive symptoms in the elderly. | This study used a prospective cohort design and two waves of panel data (1986 and 1992) from the Duke University site of the established populations for epidemiologic studies of the elderly (EPESE). | This leaves an effective sample size of 1,300, which represents all respondents who participated in the two interviews. | Functional disability. | Depression symptoms. | 6 years. | Compared to those who stayed healthy, staying disabled in Nagi or R-B is associated with a 42% increase in depressive symptoms. The proportional increases in CES-D for stable IADL and ADL disabilities are 53% and 100%, respectively. Net of all other variables, the onset of disability increases the CES-D score by 61% and this effect is highly significant. It may not be a big surprise that becoming severely disabled in ADLs results in the largest and most significant increase in depressive symptoms (80%). | The results of this study underscore the dynamics of the relationship between changing functional disability and the dynamics of depressive symptoms over time in late life. Stable disability statuses in strength and mobility, instrumental activities of daily living (IADL) items and activities of daily living (ADL) items have increasing effects on increment in CES-D scores by the follow-up. The onset of disability has stronger effects on change in CES-D scores than recovery. |
| Kivela S.L. et al., 2001 | 2001/3/1 | Cohort study. | To discover whether the occurrence of depression in community-living older people predicts impairment of physical functioning independently after adjustment for the known risk factors. | An epidemiological and clinical survey of depression in old age was performed in the town of Ahtari, western central Finland, in 1984/1985. | The actual study material consisted of 786 persons (323 men and 463 women). | Depression. | Impairment of physical functioning. | 5 years. | The occurrence of depression with a long term or relapsingcourse during follow-up (RR=1.7, 95% CI=1.01-2.71) and the onset of depression during follow-up in persons not depressed at the baseline ( RR=2.4, 95% CI=1.55-3.65) predicted lowering of functional abilities during follow-up. | Depression that developed during the follow-up in previously nondepressed persons was associated with an increased risk for lowering offunctional abilities. |
| Lisa C. Barry et al., 2009 | 2009/9/23 | Cohort study. | The authors evaluated the association between level of depressive symptoms and severity of subsequent disability over time and determined whether this relationship differed by sex. | Data from a unique longitudinal study that includes monthly assessments of disability for up to 9 years along with serial assessments of depressive symptoms at 18-month intervals. | 754 persons were included in this study. | Depressive symptoms. | Disability. | 9 years. | Moderate (odds ratio = 1.30; 95% confidence interval: 1.18–1.43) and high (odds ratio = 1.68; 95% confidence interval: 1.50–1.88) depressive symptoms were associated with mild disability, whereas only high depressive symptoms were associated with severe disability (odds ratio = 2.05; 95% confidence interval: 1.76–2.39). | Our findings indicate that depressive symptoms among older persons contribute substantially to the burden of disability over time and demonstrate the potential adverse consequences of depressive symptoms that do not reach the threshold for subsyndromal depression. |
| Johan Ormel et al., 2002 | 2002/7/1 | Cohort study. | To examine the temporal character of the reciprocal effects between IADL/ADL disability and depressive symptoms. | The article reports on a cohort of 753 persons from the Groningen Ageing Study (GLAS) who were eligible—as a result of their physical limitations—for follow-up during 2 years with three waves of measurement. | The cohort of 753 participants included from the source population only those who had four or more physical limitations. | IADL/ADL disability and depressive symptoms. | Depressive symptoms and IADL/ ADL disability. | 2 years. | Change in state disability had a moderately strong contemporaneous effect of 0.41 on depressive symptoms, but no 1-year lagged effect. Change in depressive symptom level did not have a significant immediate contemporaneous influence on disability, but it had weak 1-year lagged effect of 0.19. | The association between disability and depression could be separated into three components: (a) a strong contemporaneous effect of change in disability on depressive symptoms, (b) a weaker 1-year lagged effect of change in depressive symptoms on disability, and (c) a weak correlation between the trait (or stable) components of depression and disability. |
| F. Curtis Breslin et al., 2006 | 2006/8/1 | Cohort study. | This study examines the relationship between major depression, subclinical depressive symptomatology and activity limitation, with a particular emphasis on examining gender differences. | Data from the National Population Health Survey (NPHS). | The present analyses are based on the 7732 respondents in this age range. | Depressive symptoms. | Activities limitation. | 4 years. | The odds of respondents with a major depressive episode having an activity limitation were between 3.4 and 5.7 times as high as those reporting no depressive symptoms. | The findings provide further evidence that major depression leads to impairments in a range of daily activities. Gender differences in the impact of depression on leisure activities may be important to consider in depression treatment. |
| Luca Dalle Carbonare et al., 2009 | 2009/2/1 | Cohort study. | This study was undertaken to evaluate whether depressive symptoms predict physical disability in elderly individuals. | Data from Italian Longitudinal Study on Aging (ILSA) was used to analysis in this study. | The complete data of 3,256 subjects were collected and analyzed. | Depression symptoms. | Physical disability. | 4 years. | Incidence rates of both ADL and PPT disability were significantly associated with depressive symptoms (χ2=32.61, df=1, p<0.0001 for ADL; χ2=14.01, df=1, p<0.0002 for PPT). | Our analyses show that DS is an important predictor of self-reported physical disability among men, and the adjustment for other variables does not change OR for DS. |
| Coen H. van Gool et al., 2005 | 2005/1/1 | Cohort study. | This study determines the presence of the main pathway of disablement in a cohort aged 55 years and older and examines whether progression of the main pathway of disablement is accelerated in the presence of depression. | The Longitudinal Aging Study Amsterdam (LASA) is a cohort-study on predictors and consequences of changes in well-being and autonomy in the late middle aged and older population. | Finally, a study sample of 1110 healthy and diseased respondents. | Depression symptoms. | Disablement process. | 6 years. | For depressed respondents Functional limitations at T1 lead to almost twice as much Disability at T2 than Functional Limitations lead to Disability for non-depressed respondents (unstandardized beta regression coefficients, respectively, 0.236 and 0.121). | Depression significantly modified the associations between Pathology and subsequent Impairments, and between Functional Limitations and subsequent Disability. |
| Päivi Lampinen et al., 2003 | 2003/6/1 | Cohort study. | The present study examines the relative roles of mobility status and physical activity as predictors of depressive symptoms among community-dwelling older adults. | The data for this study were collected as part of the Evergreen Project, a prospective study on health and functional capacity among the residents of the city of Jyväskylä, central Finland, aged 65 and over. | The final sample (N=384) consisted of all the men and women who had participated in both 1988 and 1996. | Mobility and physical activity. | Depression symptoms. | 8 years. | Those who had mobility problems and were classified as sedentary (Disabled-Sedentary group) at baseline had about a 2.5-fold risk of experiencing depressive symptoms at follow-up, compared with those who had good mobility and a physically active life-style. In the Disabled-Active group, the risk also seemed to be somewhat higher (OR=1.99). | Mobility problems and older age seem to increase the risk for developing depressive symptoms in elderly people. The risk is not associated with the level of physical activity. |
| SUSAN A. EVERSON -ROSE et al., 2005 | 2005/7/1 | Cohort study. | We investigated whether depressive symptoms predicted change in physical function in elderly adults. | Participants were from the Chicago Health and Aging Project (CHAP), an ongoing longitudinal study of risk factors for incident Alzheimer disease and other age-related chronic conditions. | We limited the analyses to 4069 participants with valid CES-D data at the baseline assessment and valid physical performance data at baseline and at least 1 follow-up interview. | Depression symptoms. | Physical Function. | 6 years. | Adjusting for age, sex, race, and education, each 1-point higher CES-D score was associated with a 0.34-point lower absolute level of physical performance (p<0.0001), but there was no evidence of a CES-D by time interaction (p=0.84). Compared with the referent group (CES-D=0), the 2 middle CES-D categories (CES-D=1 or 2–3) evidenced some decline in physical performance over time, but the highest CES-D group (CES-D≥4) showed no significant physical decline over time (p<0.89). | We observed a strong cross-sectional association between depressive symptoms and overall physical performance. Physical function declined over time, yet depressive symptoms did not consistently contribute to greater decline over an average of 5.4 years of follow up among older adults. |
| S.W.GEERLINGS et al., 2001 | 2001/11/1 | Cohort study. | The longitudinal effect of depression on functional limitations and disability (in terms of disability days and bed days) was studied, thereby taking into account the role of chronic physical diseases. | The Longitudinal Aging Study Amsterdam (LASA) provides a unique resource for investigating the longitudinal relationship between depression and functional outcomes in the elderly. | The study is based on a sample which at the outset consisted of 325 non-depressed and 327 depressed persons (55–85 years). | Depression symptoms. | Functional limitations and disability. | 10 years. | Depression at baseline (CES-D≥16) was also predictive for functional limitations (incidence density rate (IDR) =2.48, 95%CI:1.89–3.27), disability days (B=0.45, SE=0.08, P< 0.001), and bed days (B=0.24,SE=0.05, P<0.001) over time. | A statistically significant association was found between depression at one point in time and both functional limitations, the number of disability days and the number of bed days 5 months later. |
| Jingmei Jiang et al., 2004 | 2004/9/1 | Cohort study. | This study examined the influence of depressive symptoms on the prevalence of physical disability and analyses the role of some confounding variables in this relationship. | Data for present study were obtained from the Beijing Longitudinal Study of Aging (BLSA). | A cohort of 1828 elderly aged 55 and older who were initially free of any physical disability. | Depression symptoms. | Physical disability. | 8 years. | Depression was found to have significantly increasing the risk for BADL disability and IADL disability. The relative risk associated with depression was stronger for IADL disability (RR: 4.98, 95%CI: 2.46, 10.09) than BADL disability (RR: 2.52, 95%CI: 2.02,4.82). When education, economic status and area of residence were considered, the risk associated depression declined to 2.48 (95%CI: 1.52, 4.08), and addition of baseline chronic conditions further reduced the risk to 2.20 (95%CI: 1.33, 3.62). Included all controlled factors further reduced the risk for IADL disability to 4.29 (95%CI: 2.08, 8.86). | This study provides the evidence that depressive symptoms are associated with significantly increased risks for subsequent BADL disability and IADL disability with noticeably higher prevalence of disability among depressed persons than non-depressed persons, and this trend remaining at each follow-up interview. |
| Joyce T. Bromberger et al., 2009 | 2009/1/1 | Review. | The purpose of the current research is to review the literature since 1966 for studies examining the association between depression and physical and psychosocial impairment in midlife women. | We include only longitudinal studies in the current review as they are the best way available to try to disentangle the relationship. PubMed, Medline, and PsychINFO were searched for articles published between January 1966 and April 2009. | 14 studies were found to meet all the inclusion criteria. | Depression and functional status. | Functional status and depression. |  |  | Results of the review indicate evidence for bi-directional associations between depression and functioning in middle-aged women. However, the studies are only broadly informative. Most adjusted for only a limited group of factors that could be associated with both depression and functioning. None of them directly examined potential moderators or mediators of the relationship between depression and impaired functioning. |
| Brenda W.J.H. Penninx et al., 2001 | 2001/1/1 | Cohort study. | The present study investigates the impact of chronicity and changes in depression on physical decline over time among community-dwelling older persons. | Data for this study were collected in the Longitudinal Aging Study Amsterdam, a prospective study of older persons aged 55–85 years. | Complete follow-up data were available for 2121 Physical function was measured by an observation subjects. | Depression. | Physical decline. | 3 years. | After adjustment for these covariates, emerging and chronic depression significantly increased the risk for substantial decline in performance (OR=1.76, 95%CI=1.23–2.51 and OR=1.67, 95%CI=1.09–2.58, respectively).When compared to never depressed persons, the adjusted risk for incident physical disability after 3 years of follow-up was significantly increased among those with emerging depression (OR=2.41, 95%CI=1.45–3.99) and chronic depression (OR=2.66, 95%CI=1.31–5.38). | Our findings among community-dwelling older persons show that chronicity of depression has a large impact on physical decline over time. |
| Jingmei JIANG et al., 2002 | 2002/1/1 | Cohort study. | The purpose of this study was to test the hypothesis that ADL disability affects the risk of onset of depressive symptoms and the role of possible confounding variables in this relation. | Data for these analyses came from the Beijing Longitudinal Study of Aging (BLSA). | The analytic cohort study for prospective analysis consisted of 1,680 subjects. | ADL disability. | Depression. | 8 years. | IADL disability was associated with a higher risk (RR=4.96) for depression than that associated with BADL disability (RR=3.56). In BADLs, three most basic items (getting in and out off bed, feeding and grooming) were strongly associated with these risks for depression (RR=7.44, 6.13, 6.07, respectively). In IADLs, the items which connected mobility (walking 300 meters, walking up and down stairs and shopping) were significantly associated with risk for depression (RR=5.44, RR=4.84 and RR=4.71, respectively). | ADL disability significantly increases the risk for subsequent depressive symptoms. At each follow-up interview, the depressive risk associated disability (in both urban and non urban areas) was significantly higher than those associated with most other sociodemographic factors. |
| Mari Kazama et al., 2011 | 2011/5/1 | Cohort study. | To determine whether symptoms of depression are associated with a subsequent decline in higher-level ADLs within a 12-month period of time. | Data from the Yamanashi Healthy Active Life Expectancy (Y-HALE) Study. | 587 non-institutionalized adults aged ≥65 years | Depression symptoms. | Higher-  Level ADLs. | 1 year. | The relative risk (RR) for a decline in higher-level ADLs in those showing signs of severe depression symptoms was 3.2 (95% CI 1.6–6.3) after adjusting for all covariates. When we evaluated the effect of severe depression symptoms on the three components of the TMIG-IC, the fully adjusted RRs (95% CIs) were 6.8 (3.1–14.8), 1.8 (0.8–4.4), and 1.9 (0.9–4.2) for declines in social role function, intellectual activities, and instrumental self-maintenance, respectively. | Severe depression symptoms were strongly associated with an increased incidence of decline in higher-level ADLs within a 12-month period of time. |
| Erin Dunne et al., 2011 | 2011/1/1 | Cohort study. | This longitudinal study examined the associations between older adults' goal adjustment capacities (goal disengagement and goal reengagement capacities), functional disability, and depressive symptoms. | This study included a heterogeneous, community-based sample of older adults who participated in the longitudinal Montreal Aging and Health Study Study. | The final sample consisted of 135 older adults. | Functional disability and goal adjustment capacities. | Depressive symptomatology. | 6 years. | The study concluded that 4-years levels of functional disability significantly predicted increases in depressive symptoms, F(1, 124) = 5.09, P = 0.03. | Depressive symptoms and functionality disability increased over time. Moreover, poor goal disengagement capacities and high levels of functional disability forecasted six-year increases in depressive symptoms. Goal disengagement buffered the association of functional disability with increases in depressive symptoms. |
| Chun-Min Chen et al., 2012 | 2012/10/1 | Cohort study. | This longitudinal study investigates the change trajectories of both depressive symptoms and disability, as well as their associations over time. | The present study adopted a prospective study design using closed cohort baseline data from an older adult Taiwanese population obtained from the Kaohsiung City government. | Participants included 442 community-dwelling older adults. | Depression symptoms and disability. | Depression symptoms and disability. | 10 years. | This model also indicates that disability significantly contributed to the onset of depressive symptoms and vice versa. The parallel latent growth curve modeling highlights that the disability intercept had significant effects on the depressive symptoms intercept (β = 0.451), as did the depressive symptoms on disability (β = 0.215). Furthermore, the disability slope had significant effects on the slope of the depressive symptoms (β = 0.435). | This study found that disability was much more likely to affect the increase in the depressive symptom trajectory over time, as compared with the influence of depressive symptoms on the increase of the disability trajectory over time. A strong association between the initial onset of disability with the initial onset of depressive symptoms and vice versa as well as between the development of both disability and depressive symptoms. |
| Chun-Te Lee et al., 2012 | 2012/11/1 | Cohort study. | The study aimed to examine the outcome predictors of improvement in depressive symptoms in the elderly over a 4-year follow-up period. | Data from the Survey of Health and Living Status of the Elderly in Taiwan (SHLSET). | The final group with complete data obtained in both 2003 and 2007 numbered 206. | Demographics, chronic medical diseases, and health-related behaviors. | Depressive symptoms. | 4 years. | Compared to those with low mobility difficulty, subjects with a high or moderate difficulty in terms of mobility limitations had ORs of depression of 0.31 (P=0.0009) and 0.47 (P=0.0326), respectively. More social support (OR=2.10, 95%CI =1.02–4.32, P=0.0442) and fewer mobility limitations (OR=0.42, 95%CI=0.19–0.93, P=0.0319) were independent significant predictors of improvement associated with depressive symptoms in the elderly. | The independent predictors related to improvement in depressive symptoms in the elderly are more social support and fewer mobility limitations. The 2 items of mobility limitations most associated with non-improvement of depression were difficulty in carrying things and squatting. |
| Ya-Ting Yang et al., 2015 | 2015/11/1 | Cohort study. | The objective of this study is to investigate the interrelationships between disability, somatic diseases and the onset of depression in seniors. | Data from the Survey of Health and Living Status of the Elderly in Taiwan (SHLSET). | The total number of 1467 participants was conducted. | Demographics, chronic medical illnesses, the change of subjects’ self-perceived health status, functional limitations and mobility limitation. | Depression. | 4 years. | Functional limitations, particularly IADL (adjusted OR = 1.81, 95% CI: 1.24–2.65, P = 0.002) and ADL (adjusted OR = 1.77, 95% CI:1.27–2.47, P = 0.001) were independently associated with the onset of depression among the elderly population. | That heart conditions and joint disorders, as well as functional limitations, particularly IADL and ADL were closely correlated with the onset of depression among the elderly population. |
| Peter A. Coventry et al., 2020 | 2020/1/1 | Cohort study. | To evaluate if depression contributes, independently and/or in interaction with frailty, to loss of independence in instrumental activities of daily living (ADL) in older adults with frailty. | Data from the Community Ageing Research 75+ Study (CARE75+). | We identified baseline data for 553 participants. | Frailty and depression. | IADL disability. | 1 year. | The coefficient for depression was also statistically significant and indicate that depressed individuals had 6.4 points (95%CI: -8.274 to -4.549) lower NEADL score compared with non-depressed individuals). The interaction between mild frailty and depression was still statistically significant (-12.5, 95%CI: -24.8 to -0.26, P<0.05). The interaction between moderate frailty and depression was weakly significant (-11.2, 95%CI: -23.1 to 0.71, P<0.1). The interaction term was not significant for severe frailty (-7.3, 95%CI: -19.5 to 5.0). | Frailty and depression are independently associated with reduced independence in instrumental activities of daily living. Also, depression interacts with frailty to further reduce independence for mild to moderately frail individuals. |
| Kathryn L. Bacon et al., 2016 | 2016/2/9 | Cohort study. | The current study evaluated the longitudinal and reciprocal associations between depressive symptoms and disability over 6 years, and whether these associations differed in caregivers and noncaregivers. | The sample used for this analysis was derived from Caregiver Study of Osteoporotic Fractures (SOF). | The sample was restricted to 956 participants. | Depression symptoms and disability. | Depression symptoms and disability. | 6 years. | Higher depressive symptoms predicted greater disability at the following interview (beta = 0.02, P<0.05), and greater baseline disability also predicted higher depressive symptoms at the following interview (beta = 0.54, P<0.05).In contrast to noncaregivers, depressive symptoms and disability were not significantly associated over time among caregivers (beta=0.01, P=0.80, relating depressive symptoms at baseline to disability at the second follow-up interview). | This study found that depressive symptoms and disability formed reciprocal longitudinal relationships in older women noncaregivers, but not caregivers followed for 6 years. |
| Takahiro Nakamura et al., 2017 | 2017/12/1 | Cohort study. | The objective of the study was to examine the association between depressive symptoms and future ADL dependence and to investigate how this association varies according to living circumstances and marital status. | Data from the Kurabuchi Study. | The total study population consisted of 769 participants (318 men, 451 women). | Depressive symptoms. | Dependence in Activities of Daily Living (ADLs). | 7.5 years. | Those with depressive symptoms were more prone to future dependence in ADLs, even after adjustment for potential confounders (aRR=1.29, 95% CI=1.04–1.61). | Depressive symptoms are associated with future ADL dependence and that living circumstances (except for institutionalization) and marital status do not affect the association. |
| Jin-Won Noh et al., 2016 | 2016/11/30 | Cohort study. | The purpose of this study was to examine the relationship between physical disability and depression by gender among adults. | Data from the Korean Longitudinal Study of Ageing (KLoSA). | All 10,254 participants were included. | Physical disability. | Depression. | 6 years. | Those who were female [B (SE)=0.21 (0.08), P =0.008] or diagnosed with physical disability by doctors [B (SE)=1.05 (0.13), P<0.001] presented higher scores in level of depression. | This study, using a nationally representative sample, shows that being female and disabled is associated with depression. |
| Kathrin Heser et al., 2018 | 2018/6/26 | Cohort study. | This study examines the relationship between late-life depressive symptoms, cognitive and functional impairment in a cohort of very old community-based participants. | Data from the German AgeMooDe study. | A sample of 1,226 primary care patients was included. | Depressive symptoms and functional impairment. | Depressive symptoms and functional impairment. | 1 year. | An increased risk of subsequent functional impairment at follow-up was associated with more depressive symptoms at baseline (OR=1.25, 95%CI=1.15–1.37, P<0.001) and with an increase of depressive symptoms from baseline to follow-up (OR=1.28, 95%CI=1.13-1.44, P<0.001). A decline of functional abilities from baseline to follow-up predicted an increased risk for elevated depressive symptoms at follow-up (OR=1.19, 95%CI=1.11–1.27,P<0.001) | Depressive symptoms and global cognitive function were not associated longitudinally, but level and increase of depressive symptoms over time predicted functional impairment after 1 year. |
| He Minfu et al., 2018 | 2018/8/12 | Cohort study. | To describe the prevalence of depression symptoms among people with different degrees of ADL disability and their spouses and evaluate the association between baseline ADL disability and development of personal and spouse depression symptoms. | Data derived from the China Health and Retirement Longitudinal Study (CHARLS). | There were 15,890 subjects included in the study, | ADL disability. | Depression symptoms. | 2 years. | Prospectively, BADL score ≥2 was associated with higher risk of depression symptoms of subjects (OR 1.63, 95%CI 1.03–2.57) and their spouses (OR 1.50, 95%CI 1.01–2.22). | Among middle-aged and older Chinese adults, functional impairment in ADL was associated with increased risk of depression symptoms not only in the disabled themselves but also their spouses without ADL disability. |
| Mengxiao Hu et al., 2023 | 2023/11/24 | Cohort study. | This study aimed to quantify the longitudinal mediating effect of ADL disability in the association between the accumulation of chronic conditions and depressive symptoms in both the United States and China. | This population-based cohort study used data from the Health and Retirement Study and the China Health and Retirement Longitudinal Survey. | A total of 22,335 middle-aged and older adult were included. | Chronic conditions and depressive symptoms. | Chronic conditions and depressive symptoms. | 4 years. | Respondents with disability (T2) were more likely to have more depressive symptoms (T3) (men: β=0.105, [95%CI, 0.078–0.131], P<0.001; women:β=0.157, [95%CI, 0.135–0.180]). Moreover, depressive symptoms (T2) predicted disability in the subsequent wave (men: β=0.088, [95%CI, 0.067–0.109]; women: β=0.057, [95%CI, 0.041–0.073]). The accumulation of chronic conditions (T1) was positively associated with ADL disability (T2) (path a: men: β=0.081, [95%CI, 0.063–0.099]; women: β=0.087, [95%CI, 0.072–0.101]), and ADL disability (T2) were also related to increased depressive symptoms (T3) (path b: men: β = 0.105, [95%CI, 0.078–0.131]; women: β=0.157, [95%CI, 0.135–0.180]). | This study revealed the longitudinal mediating effect of ADL disability on the association between the accumulation of chronic diseases and depressive symptoms. The study documented the bidirectional association among the accumulation of chronic conditions, ADL disability, and depression in both the United States and China. And the mediation proportion was greater for women than for men. |
| Dexia Kong et al., 2019 | 2019/8/1 | Cohort study. | This prospective cohort study examined the relationship between depressive symptoms and onset of functional disability over 2 years among US Chinese older adults. | Data were obtained from the Population Study of Chinese Elderly in Chicago (PINE). | The present study used data from 2713 participants who completed both the baseline and follow-up interviews. | Depressive symptoms. | Functional Disability. | 2 years. | Odds of ADL disability onset (OR=1.06; 95% confidence interval [CI]=1.02-1.11), IADL disability onset (OR=1.05; 95%CI=1.01-1.09), and mobility disability onset (OR=1.05; 95%CI =1.01-1.09) were consistently higher in US Chinese older adults with higher levels of depressive symptoms than their less-depressed counterparts. | Findings indicate that depressive symptoms predict subsequent onset of ADL, IADL, and mobility disabilities over 2 years among US Chinese older adults. |
| Juliana Lustosa Torres et al., 2018 | 2018/1/26 | Cohort study. | To examine the ability of a social support, social network and depressive symptoms baseline measures to predict onset of ADL disability in long term in a Western middle income country. | Data from the Bambuí (Brazil) Cohort Study of Aging. | 1,014 participants aged 60 years and older were included. | Depressive symptoms. | ADL disability. | 15 years. | Regarding those with no depressive symptoms and high support, low emotional support and depressive symptoms alone increased the risk of disability (SHR = 1.11; 95%CI: 1.01; 1.45 and SHR=1.52; 95%CI: 1.13; 2.01, respectively). The presence of both factors increased the risk of disability by 1.61 (95%CI: 1.18; 2.18). | Baseline measures of both depressive symptoms and emotional support, have predictive value for incident disability in long term. |
| Gina Lee et al., 2023 | 2023/1/31 | Cohort study. | The purpose of the study was to examine a bivariate latent change score model of depressive symptoms and functional limitations (activities of daily living) among centenarian or near-centenarian survivors. | Data from the Health and Retirement Study (HRS). | 460 participants who eventually survived to age 98 or older were included. | Depressive symptoms and functional limitations. | Depressive symptoms and functional limitations. | 6 years. | Parameter estimates of Model 3 suggest significant paths from depressive symptoms to subsequent changes in depressive symptoms (β=−1.307, P<0.001), which indicates lower number of depressive symptoms is associated with an increment in the number of depressive symptoms at subsequent time points. Also, significant paths from depressive symptoms impacting subsequent changes in functional limitations were identified (β=3.793, P<0.05). | Conclusively, the findings of our study reveal that depressive symptoms was the leading variable in the relationships between depressive symptoms and functional limitations among centenarian survivors when they were in their 80s. |
| Ulrike Dapp et al., 2020 | 2020/11/6 | Cohort study. | To investigate the mutual relationships between frailty, disability and depression/depressed mood. | Data were obtained from the Longitudinal Urban Cohort Ageing Study (LUCAS) in Hamburg, Germany. | The numbers of participants of 2012 were inccluded. | Depressed mood, functional decline and disability. | Depressed mood, functional decline and disability. | 10 years. | Depressed mood significantly increased the hazard of subsequent functional decline (HR=1.581; 95%CI: 1.257 to 1.988; P<0.001). Functional decline significantly increased the hazard of subsequent depressed mood (HR=2.324; 95%CI: 1.703 to 3.172; P<0.001). Depressed mood significantly increased the hazard of subsequent disability (HR=2.589; 95%CI: 1.885 to 3.557; P<0.001). Disability did not significantly increase the hazard of subsequent depressed mood (HR=1.540; 95%CI: 0.917 to 2.579; P=0.102). | Depressed mood predicted subsequent occurrence of frailty and, even stronger, subsequent BADL dependency. Conversely, frailty predicted subsequent depressed mood but BADL dependency did not significantly predict later depressed mood. |
| Rumei Yang et al., 2021 | 2021/7/3 | Cohort study. | To examine whether, and to what degree, the rate of change in physical functioning over time was associated with depressive symptoms, subjective memory and cognitive functioning. | Data from the China Health and Retirement Longitudinal Study. | The sample included 5,519 older adults. | Depressive symptoms and cognitive performance. | Physical functioning. | 4 years. | Compared with those with low depressive symptoms (CESD<10), older adults with high depressive symptoms (CESD≥10) had significantly higher levels of mobility (β =0.61, 95%CI: [0.48, 0.74], P < 0.001), ADLs (β=0.26, 95%CI: [0.19, 0.34], P<0.001) and IADLs (β=0.28, 95%CI: [0.21, 0.36], P<0.001) impairment. | In our sample, high depressive symptoms were associated with accelerated mobility, ADLs and IADLs impairment, and poor cognitive performance was associated with accelerated mobility impairment. |
| Hongting Ning et al., 2021 | 2021/9/21 | Cohort study. | To determine the impact of baseline metabolic syndrome and depressive symptoms on subsequent functional disability. | Data from the 2011 baseline and 2013, 2015 and 2018 follow-up waves of the China Health and Retirement Longitudinal Study (CHARLS). | A total of 5475 participants were included in the current study. | Metabolic syndrome and depressive symptoms. | Functional ability. | 7 years. | Baseline depressive symptoms significantly predicted functional disability over a 7-year follow-up after adjusting for covariates (Hazard ratio [HR]=1.54, 95% confidence intervals [CI]=1.40–1.70 for ADL disability; HR=1.36, 95%CI=1.25–1.48 for IADL disability). | The current study found that baseline depressive symptoms were significantly associated with both ADL and IADL disabilities, while metabolic syndrome significantly predicted ADL disability. |
| Fan Tian et al., 2022 | 2022/5/2 | Cohort study. | We prospectively examined the association of functional disability with trajectories of depressive symptoms over time among middle-aged and elderly Chinese adults in a large population-based cohort. | Data were drawn from the China Health and Retirement Longitudinal Study. | A total of 8415 participants were included. | Functional disability. | Depressive symptoms. | 7 years. | Participants with severe functional disability were at increased likelihood of being in the moderate (odds ratio [OR]=2.27, 95% confidence interval [CI] 1.68–3.07), increasing (OR=2.31, 95%CI 1.49–3.59), and high (OR =4.74, 95%CI 3.07–7.31) depressive symptom trajectories. | Our findings suggest that functional disability was associated with unfavorable depressive symptom trajectories among middle-aged and older Chinese adults. |
| Jiayi Wang et al., 2023 | 2023/6/11 | Cohort study. | The study’s objective was to assess the reciprocal relationship between ADL disability and depressive symptoms among middle-aged and older Chinese people. | Data was collected in the China Health and Retirement Longitudinal Study (CHARLS). | This study included 4,124 participants aged ≥ 45 years. | Activities of daily living disability and depressive symptoms. | Activities of daily living disability and depressive symptoms. | 7 years. | The paths d1 (β=0.070, SE=0.015, P<0.001), d2 (β = 0.069, SE=0.014, P<0.001), d3(β=0.053, SE=0.014, P<0.001) from ADL disability to depressive symptoms between 2011 and 2018 were statistically significant. There were positive paths e1 (β=0.106, SE=0.015, P<0.001), e2 (β=0.099, SE=0.014, P<0.001), e3 (β = 0.073, SE=0.014, P<0.001) from depressive symptoms to ADL disability. | This study shows that ADL disability is bi-directionally related to depressive symptoms in middle-aged and older Chinese people over time. |
| Weihao Wang et al., 2024 | 2024/1/27 | Cohort study. | To determine the effect of various patterns of functional disability and new-onset functional disability on depressive symptoms among Chinese older adults aged 60 years and above. | Data from the China Health and Retirement Longitudinal Study (CHARLS), | The study included 3242 older adults. | Depressive symptoms. | Functional Disability. | 7 years. | Participants with IADLs disability alone (HR=1.29; 95%CI: 1.05–1.58) and with both IADLs and BADLs disabilities (HR=1.78; 95%CI: 1.41–2.24) had an increased risk of depressive symptoms. There was no statistically significant association between BADLs disability alone and depressive symptoms (HR=1.22 95%CI: 0.96–1.55). | Functional disability increases the risk of depressive symptoms, particularly impaired IADLs function. |
| Eric J. Lenze et al, 2005 | 2005/3/30 | Cohort study. | To examine the relationship between persistently high depressive symptoms and long-term changes in functional disability in elderly persons. | Participant data from the Cardiovascular Health Study. | The CHS enrolled 5,888 men and women in four communities. | Depression. | Functional Disability. | 4 years. | The persistently depressed group had an adjusted odds ratio (OR) of 5.27 (95% confidence interval (CI 3.03–9.16) for increased functional disability compared with the nondepressed group over 3 years of follow-up, whereas the temporarily depressed group had an adjusted OR of 2.39 (95%CI 5 1.55–3.69) compared with the nondepressed group. | Persistently elevated depressive symptoms in elderly persons are associated with a steep trajectory of worsening functional disability. |
| Lydia W. Li et al, 2008 | 2008/6/25 | Cohort study. | To investigate the effect of changes in depression status on physical disability in older persons receiving home care, examine whether the effect is due to concomitant changes in cognitive status, and test whether affective state and cognitive ability interact to influence physical disability. | Data collected from a large cohort of elderly participants in two publicly funded home- and community-based long-term care programs in Michigan: Medicaid Waiver and Care Management. | The final sample consisted of 13,129 respondents. | Depressive symptoms and cognitive functioning. | Physical disability. | 5 years. | A change from subthreshold to no depression is associated with a decrease of 0.104 point (SE=0.033, p=0.001) in ADL score. An improvement from subthreshold to no depression is significantly associated with a subsequent decrease in IADL score both before(beta=−0.040, SE=0.011, p<0.001) and after (beta=− 0.036, SE=0.011, p=0.002) adjusting for cognitive status. A deterioration from borderline intact to impaired cognition is associated with an increment of 0.251 point (SE=0.051, p<0.001) in ADL score and 0.099 point (SE=0.020, p<0.001) in IADL score. The interaction of depression and cognitive status on IADL scale score is statistically significant. | This analysis shows that worsening of depression and cognitive status independently predicts a subsequent increase in ADL and IADL score (more disability) and an in creased likelihood of dependency in both domains of functioning among home care elders. Transitions between no and subthreshold depression and between borderline intact and impaired cognition are particularly important to their change in physical disability. |
| Isabelle Carrière et al, 2011 | 2011/3/3 | Cohort study. | To examine the long-term association between depressive symptoms and incident activity limitations in a large elderly prospective community cohort, for which information on a large number of potential confounding factors was available. | Subjects were recruited as part of a multi-site cohort study of community-dwelling persons aged 65 years and over from the electoral rolls of three French cities (Bordeaux, Dijon and  Montpellier) between 1999 and 2001. | The present analyses were thus conducted on 3191 subjects. | Depression. | Activity limitations. | 7 years. | In men, mild depressive symptomatology was associated with increased incident limitations on instrumental activities of daily living (IADL) (odds ratio (OR) (95%CI)=5.07(2.25-11.42)). In women, severe depressive symptomatology was related to social restriction (OR(95%CI)=2.36(1.31-4.25)), IADL (OR(95%CI)=1.89(1.13-3.15)) and activities of daily living (OR(95%CI)=11.15(3.43-36.23)). | The relationship between DS and incident activity limitations in the elderly is gender-dependent and also varies according to symptom load. |
| Xia Li et al, 2012 | 2012/8/15 | Cohort study. | The study was to assess distinctive patterns for the development of physical limitation and depression and to explore their correlation to form a proper prevention strategy. | Data from the Beijing Longitudinal Study of Aging (BLSA) 1992–2009 hosted by Xuanwu hospital. | 316 subjects were engaged in this analysis. | Physical limitation. | Depression. | 10 years. | When gender, age and number of chronic conditions were adjusted, IADL trajectory group was an independent risk factor for the adverse CES-D trajectory group. Subjects from the Late increase group in IADL scores were more likely found in the adverse CES-D group, with ORs of 3.900 (95%CI: 1.347–11.290). | Given the important impact of activities of daily living functioning on utilization of medical services and quality of life, prevention or reduction of depressive symptoms should be considered as an important reason for intervention. |
| Mary Elizabeth Bowen et al, 2015 | 2015/5/7 | Cohort study. | To examine how the relationship between depressive symptoms and disability may vary by nativity status in later life. | Data from the Health and Retirement Study (HRS). | The study included 15444 older adults. | Depression. | Disability. | 12 years. | Depressive symptoms were associated with increased IADL and ADL disability among Latinos compared with Whites; foreign-born Latinos had lower than expected depressive symptom–related IADL and ADL (0.82; p ≤ .001) disability. | This prospective study of older adults in the United States found that foreign born Latinos have lower than expected depressive symptom–related IADL and ADL disability than their older White and U.S.-born Latino counterparts. |
| Anda Botoseneanu et al., 2023 | 2023/6/20 | Cohort study. | To evaluate the impact of depressive multimorbidity on the long-term development of activities of daily living (ADL) and instrumental activities of daily living (IADL) limitations according to racial/ethnic group in a representative sample of US older adults. | Data from the Health & Retirement Study (HRS). | The final analytic sample consisted of 16,364 self-respondents. | Depressive multimorbidity. | Activities of daily living (ADL) and instrumental Activities of Daily Living (IADL). | 16 years. | Depressive and somatic multimorbidity were associated with 5.18 and 2.95 times greater accumulation of functional limitations, respectively, relative to no disease (incidence rate ratio [IRR]=5.18, 95%CI[4.38,6.13], IRR=2.95, 95%CI[2.51,3.48]). | Combinations of somatic diseases and high depressive symptoms are associated with greatest accumulation of functional limitations over time in adults ages 65 and older. |
| **The relationship between sleep and depression** | | | | | | | | | | |
| Yankun Sun et al., 2018 | 3/31/2018 | Cohort study. | To identify the prospective effect of sleep duration on the incidence and recurrence of depression, as well as the influence of depression on changes of sleep patterns in community-dwelling mid-age and elderly individuals. | Data from the China Health and Retirement Longitudinal Study (CHARLS). | 10,704 participant were included for baseline and four-year follow up. | Sleep duation. | Depression. | 4 years. | Participants with short sleep duration (<5 and 5-6 h) had a higher risk of depression onset (OR 1.69 [1.36-2.11], 1.48 [1.19-1.84]) and recurrent depression (OR 1.44 [1.12-1.86], 1.32 [1.00-1.74]) compared to participants with normal sleep durations (7-8 h). Individuals with depression were more likely to have short sleep durations instead of long ones (RRR 1.20 [1.02-1.43]). | The present study identified the bidirectional relationship between sleep duration and depression. Short sleep duration was associated with an increased risk of onset and recurrent depression. Depression will shorten the sleep duration of individuals with normal sleep durations rather than extend it. |
| Lydia Poole et al., 2017 | 11/4/2017 | Cohort study. | The reasons for the comorbidity between depressed mood and poor sleep are not well understood. | Data from the English Longitudinal Study of Ageing (ELSA). | Participants were 5172 adults aged 50 years and older. | Depressive symptoms and sleep symptoms. | Depressive symptoms and sleep symptoms. | 4 years. | Analyses predicting wave 6 depressive symptoms showed significant predictors to greater baseline sleep complaints (β = 0.067, 95%CI 0.135–0.311), and the increased odds of sleep complaints at wave 6 showed baseline depressive symptoms (OR 6.795, 95%CI 5.860–7.879) to be a significant predictor. | Depressive symptoms and sleep complaints share a range of correlates cross-sectionally and prospectively. |
| Hyong Jin Cho et al., 2008 | 1/1/2008 | Cohort study. | To test whether sleep disturbance predicts depression recurrence independent of other depressive symptoms. | A 2-year prospective cohort study was conducted with community-dwelling older adults ages 60 years and older. The participants were assessed at baseline, 6 weeks, 1 year, and 2 years for depressive episodes, depressive symptoms, sleep quality, and chronic medical disease. | The study was conducted with 351 community-dwelling older adults. | Sleep Disturbance. | Depression Recurrence. | 2 years. | Depression occurrence was independently associated with group status (prior depression versus control; adjusted odds ratio=38.65, 95%CI=4.72 to 316.43, p=0.001), and also with sleep disturbance (odds ratio=3.05, CI=1.07 to 8.75, p=0.04) and other depressive symptoms (odds ratio=1.19, CI=1.07–1.33, p=0.002). Using survival analysis for time to depression recurrence, sleep disturbance was a powerful predictor of depression recurrence in the prior depression group (Mantel-Cox χ 2 =13.36, df=1, p<0.001). | This study demonstrate that sleep disturbance acts as an independent risk factor for depression recurrence in community-dwelling older adults. To identify older adults at risk for depression, a two-step strategy can be employed, which involves assessment of the presence of a prior depressive episode along with sleep disturbance. |
| Isabelle Jaussen et al., 2011 | 8/1/2011 | Cohort study. | To examine the relationship between sleep disturbance and the incidence of daytime sleepiness over 4 years in community-dwelling elderly, taking into account sleep disturbance characteristics as well as sleep medication. | Subjects included in the present study were recruited as part of the Three City Study, an ongoing multi-site prospective study involving 3 French cities: Bordeaux, Dijon, and Montpellier. Briefly, 9294 subjects ≥ 65 years were recruited from the electoral rolls between March 1999 and March 2001. | The present analyses were thus performed on 3824 subjects. | Insomnia and Daytime Sleepiness. | Depressive Symptoms. | 4 years. | Insomnia symptoms and daytime sleepless independently increased the risk of incident depressive symptoms (OR=1.23, 95%CI=1.01-1.49 and OR=2.05, 95%CI=1.30-3.23, respectively). Poor sleep quality and difficulty in initiating and in maintaining sleep—but not early morning awakening—were identified as risk factors of depressive symptoms, with risk increasing with the frequency of insomnia symptoms. Sleep medication was not only a risk factor for depressive symptoms independent of insomnia symptoms (OR=1.62, 95%CI=1.26-2.09), but also independent of EDS (OR=1.71 95%=1.33-2.20). | Insomnia symptoms, daytime sleepless, and the use of medication independently increase the risk of subsequent depression in the elderly. |
| Jae-Min Kim et al., 2009 | 9/1/2009 | Cohort study. | To investigate prevalence, incidence, and persistence of insomnia, and their bidirectional longitudinal associations with depression and physical disorders. | Data from a prospective community-based study of late-life psychiatric morbidity carried out in Kwangju, South Korea from 2001 to 2003, in collaboration with the 10/66 International Research Program on dementia and late-life mental disorder in low and middle income nations. | 1204 people ≥ 65 years of age were evaluated at baseline; 909 of them (75%) were re-interviewed after 2 years. | Insomnia. | Depression and physical disorders. | 2 years. | Baseline insomnia was significantly associated with prevalent and incident depression, with an increase in number of depression symptoms, and an increase in number of reported physical disorders before and after adjustment for sociodemographic and clinical characteristics. Persistence of insomnia was strongly associated with incidence of depression over the same period (OR [95%CI]=2.4[1.3–4.2]) after adjustment. | Insomnia was common and often persistent in this population. Insomnia was closely and reciprocally related to depression and physical disorders. |
| Jeanne E Maglione et al., 2014 | 7/1/2014 | Cohort study. | To investigate the longitudinal relationship between subjective and objective sleep disturbance and depressive symptoms. | Participants were women enrolled in the Study of Osteoporotic Fractures (SOF), an ongoing, multicenter, prospective cohort study of primarily Caucasian, community-dwelling women age 65 y and older from four geographic areas. | The current analyses were performed on this subset of 952 women. | Subjective and Objective Sleep Disturbance. | Depression. | 5 years | There was an independent association between greater PSQI score at baseline and greater odds of worsening depressive symptoms (≥ 2-point increase in GDS) (Multivariate Odds Ratio [MOR] 1.19, confidence interval [CI] 1.01-1.40, P = 0.036). Higher scores specifically on the sleep quality (MOR 1.41, CI 1.13-1.77, P<0.003) and sleep latency (MOR 1.21, CI 1.03-1.41, P=0.018) PSQI subscales were also associated with greater odds for worsening depressive symptoms. Objective assessments revealed an association between baseline prolonged wake after sleep onset (WASO ≥ 60 min) and worsening depressive symptoms at follow-up (MOR 1.36, CI 1.01-1.84, P=0.046). | In older women with few or no depressive symptoms at baseline, those with more subjectively reported sleep disturbance and more objectively assessed fragmentation of sleep at baseline had greater odds of worsening depressive symptoms 5 y later. |
| Lena Mallon et al., 2000 | 9/1/2000 | Cohort study. | To investigate the natural history of insomnia and its association with depression and mortality. | From the population register, 2,663 men and women aged 45-65 years were ran-domly selected from the County of Dalarna in Sweden in 1983. The questionnaire in the follow-up survey in 1995. | 1,244 were included in this study. | Insomnia. | Depression and mortality. | 12 years. | Insomnia in women predicted subsequent depression (odds ratio [OR]= 4.1; 95% confidence interval [CI] 2.1-7.2) but was not related to mortality. In men, insomnia predicted mortality (OR=1.7; 95% CI 1.2-2.3), but after adjustment for an array of possible risk factors, this association was no longer significant. Men with depression at baseline had an adjusted total death rate that was 1.9 times hgher than in the nondepressed men (95%CI: 1.2-3.0). | In women, insomnia is a risk factor for subsequent depression, but it is not related to mortality. In men, insomnia is a risk factor for excess mortality in univariate analyses, but after adjustment for a broad array of possible risk factors in multivariate analyses, the association was no longer significant. |
| Misti Paudel et al., 2013 | 1/1/2013 | Cohort study. | Self-reported sleep disturbances are associated with an increased risk of depression in younger and older adults, but associations between objective assessments of sleep/wake disturbances via wrist actigraphy and risk of depression are unknown. | Data from the prospective Osteoporotic Fractures in Men (MrOS) Study. | 2,510 community-dwelling, nondepressed men enrolled in the MrOS Sleep study. | Sleep disturbances. | Depression. | 3.4 years. | An association between poor self-reported sleep quality and higher odds of being depressed at follow-up (multivariable odds ratio [MOR]=1.53, 95%confidence interval (CI) 1.00-2.33). In age- and site-adjusted models, objectively measured reduced sleep efficiency (odds ratio [OR]=1.88, 95%CI 1.13-3.13), prolonged sleep latency (OR=1.77, 95%CI 1.04-3.00), greater nighttime wakefulness (OR=1.48, 95%CI 1.01-2.18) and multiple long-wake episodes (OR=1.69, 95%CI 1.15-2.47) were associated with increased odds of depression at follow-up, but these associations were attenuated and no longer significant after further adjustment for number of depressive symptoms at baseline. | Among nondepressed older men, poor self-reported sleep quality was associated with increased odds of depression several years later. Associations between objectively measured sleep disturbances (e.g., reduced sleep efficiency, prolonged sleep latency, greater nighttime wakefulness, and greater long-wake episodes) and depression several years later were largely explained by a greater burden of depressive symptoms at baseline. |
| Josine G. van Mill et al., 2013 | 11/26/2013 | Cohort study. | To examine the predictive role of insomnia and sleep duration on the 2-year course of depressive and anxiety disorders. | Data were analyzed from the baseline and 2-year assessment of the Netherlands Study of Depression and Anxiety (NESDA). | A final sample size is 1,069. | Insomnia and sleep duration. | Depressive and anxiety disorders. | 2 years. | Long sleep duration was independently associated with persistence of depression/anxiety even after adjusting for severity of psychiatric symptoms (OR=2.52; 95%CI, 1.27–4.99). For short sleep duration, the independent association with persistence of combined depression/anxiety showed a trend toward significance (OR=1.32; 95%CI, 0.98–1.78), and a significant association for the persistence of depressive disorders (OR=1.49; 95%CI, 1.11–2.00). Both short and long sleep duration were independently associated with a chronic course trajectory (short sleep: OR=1.50; 95%CI, 1.04–2.16; long sleep: OR=2.91, 95%CI, 1.22–6.93). | Both short and long sleep duration—but not insomnia—are important predictors of a chronic course, independent of symptom severity. |
| Marta Jackowska et al., 2017 | 9/1/2017 | Cohort study. | This study investigated whether sleep problems, sleep duration and a combination of short or long sleep with sleep problems were predictive of depressive symptoms 6 years later. | Data from the English Longitudinal Study of Ageing. | Participants were 4545 men and women. | Sleep problems, short sleep and a combination of both. | Depressive symptoms. | 6 years. | Sleep problems were predictive of elevated depressive symptoms at follow-up (odds ratio [OR]=1.36, 95% confidence interval [CI]=1.19-1.56). When explored separately, waking up in the morning feeling tired (OR=1.71, 95%CI=1.24-2.37) followed by difficulties falling asleep (OR = 1.49, 95%CI= 1.06- 2.11) were also predictors of future depressive symptoms. Compared to optimal duration, short (OR=1.90, 95%CI= 1.34- 2.71) but not long sleep hours were also linked to elevated depressive symptoms. Participants reporting short sleep hours combined with high sleep problems also had an elevated risk of depressive symptoms 6 years later (OR=1.85, 95%CI= 1.15-3.00). | Short and disturbed sleep as well as their combination increase the risk of future depressive symptoms in older adults. |
| Rize Jing et al., 2020 | 1/1/2020 | Cohort study. | This study aimed to evaluate the longitudinal association between sleep duration and depressive symptoms among the elderly in China. | A data set from China Health and Retirement Longitudinal Study (CHARLS) in 2011, 2013 and 2015. | The final study sample included a total of 22,847 respondents. | Sleep duration. | Depressive symptoms. | 4 years. | An extra hour of total sleep including nighttime sleep and daytime nap was associated with lower incidence of depressive symptoms among the elderly after adjusting all confounders (OR=0.83, 95%CI: 0.82–0.84). In addition, an extra hour of nighttime sleep (OR=0.82, 95%CI: 0.80–0.83) or daytime nap (OR=0.93, 95%CI: 0.89–0.97) was also negatively associated with depressive symptoms among the elderly. After controlling the total sleep time, an extra hour of nighttime sleep was negatively associated with depressive symptoms (OR=0.88, 95%CI: 0.84 to 0.92), while an extra hour of daytime nap displayed a positive association with depressive symptoms (OR=0.88, 95%CI: 0.84 to 0.92). Compared with the moderate nappers, only extended nappers had significantly higher incidence of depressive symptoms (OR=1.32, 95%CI: 1.19 to 1.45). | For the elderly in China, increasing their total sleep, nighttime sleep, and/or daytime nap duration would reduce the incidence of depressive symptoms. |
| Michael J. Li et al., 2018 | 8/2/2018 | Cohort study. | The aim of this analysis was to test if changes in insomnia symptoms and global sleep quality are associated with coinciding changes in depressed mood among older adults. | We report on results yielded from secondary analysis of longitudinal data from a clinical trial of older adults. | Our sample consisted of 49 older adults. | Insomnia. | Depression symptoms. | 10 weeks. | Change in AIS scores was associated with change in BDI-II scores (β=0.38, p<0.01). Change in PSQI scores was not significantly associated with change in BDI-II scores (β=0.17, p=0.26). | Our findings suggest that improvements over ten weeks in insomnia symptoms rather than global sleep quality coincide with improvement in depressed mood among older adults. |
| Yujie Li et al., 2017 | 9/18/2017 | Cohort study. | This study aimed to evaluate the associations of nighttime sleep duration and midday napping with risk of depressive symptoms incidence and persistence among middle-aged and older Chinese. | The data analyzed in this study were derived from the China Health and Retirement Longitudinal Study (CHARLS). | There were 11,052 individuals included in the final sample. | Sleep duration. | Depressive symptoms. | 2 years. | After controlling for potential covariates, nighttime sleep duration <6 hours was associated with high risk of incident depressive symptoms (OR=1.450, 95%CI: 1.193, 1.764 for middle aged population, and OR=2.084, 95%CI:1.479, 2.936 for elderly) and persistent depressive symptoms (OR=1.404, 95%CI: 1.161, 1.699 for middle aged population, and OR=1.365, 95%CI: 0.979, 1.904 for elderly). For depressed individuals, longer midday napping (≥60minutes) was associated with lower persistent depressive symptoms (OR=0.842, 95%CI: 0.717, 0.989). | Our study concluded that short nighttime sleep duration was an independent risk factor of depressive symptoms incidence and persistence. Depressed individuals with long midday napping were more likely to achieve reversion than those who have no siesta habit. |
| Annelies Brouwer et al., 2022 | 7/1/2022 | Cohort study. | We tested whether insomnia symptoms of an older individual are associated with later depressive symptoms in that older individual, and vice versa. | Data from the Longitudinal Aging Study Amsterdam (LASA). | This resulted in a total sample‐size of 3081. | Insomnia. | Depression. | 20 years. | Symptoms of depression were not found to be associated with an additional risk of higher symptoms of insomnia three years later, and vice versa (p=0.329 and p=0.919, respectively). Similar results were found when analyses were corrected for covariates. | In older individuals, depression and insomnia are associated and tend to increase concurrently over time, but constitute no additional risk for one another over repeated three-year intervals. These findings contradict previous research that suggests that depression and insomnia are risk factors for one another over time. |
| Jialu Jiang et al., 2024 | 5/1/2024 | Cohort study. | To explore the association between sleep architecture and depressive symptoms. | Data from the Sleep Heart Health Study (SHHS). | 3247 participants were included. | Sleep architecture. | Depressive symptoms. | 5.3 years. | A significant linear association between NREM Stage 1 and depressive symptoms was found after adjusting for potential covariates. Multivariable logistic regression analysis showed that percentage in NREM Stage 1 was associated with the incidence of depressive symptoms (odds ratio [OR], 1.06; 95% confidence interval [CI], 1.02–1.10; P=0.001), as were time in NREM Stage 1 and depressive symptoms (OR, 1.02; 95% CI, 1.01–1.03; P=0.001). However, no significant association with depressive symptoms was found for other sleep stage. | Increased time or percentage in NREM Stage 1 was associated with a higher risk of developing depressive symptoms. |
| Xin-lin Li et al., 2023 | 3/2/2023 | Review with meta-analysis. | To examine the dose–response associations between night-sleep duration and depression risk in middle-aged and older adults. | Six cohort studies with 33,595 participants were included in this meta-analysis. | 33,595 participants. | Sleep duration. | Depression. | NA | On one hand, compared with 7-h of night sleep, both shorter and longer sleep duration were associated with an increased risk of depression (5 h: risk ratio=1.09, 95% confidence interval=1.07–1.12; 6 h: RR=1.03, 95%CI=1.02–1.04; 8 h: RR=1.10, 95%CI=1.05–1.15; 9 h: RR=1.31, 95%CI=1.17– 1.47; 10 h: RR=1.59, 95%CI=1.31–1.92; non-linear test p<0.05). On the other hand, an increased risk of depression with shorter sleep duration was observed in middle-aged and older people among the non Asian population (5 h: RR=1.09; 95%CI =1.02–1.17), while both shorter and longer sleep duration can increase the risk of depression among an Asian population (5 h:RR=1.10, 95%CI=1.07–1.13; 6 h: RR=1.04, 95%CI = 1.02–1.05; 8 h: RR=1.09, 95%CI = 1.05–1.14; 9 h: RR=1.35, 95%CI=1.18–1.53; 10 h: RR=1.70, 95%CI = 1.36–2.12). | The lowest-risk onset of depression occurred among middle-aged and older people with 7 h of night sleep, which suggested that shorter and longer night-sleep duration might lead to an increased incidence of depression. |
| Pengpid S et al, 2022 | 10/25/2022 | Cohort study. | To assess the relationship between sleep duration and incident depressive symptoms (IDS) and persistent depressive symptoms (PDS). | Data from the Health and Ageing in Africa: A Longitudinal Study of an INDEPTH Community in South Africa (HAALSI). | 3891 adults (≥40 years). | Sleep duration. | Depressive symptoms. | 4 years. | Long sleep duration was positively associated with IDS among men (AOR: 1.37, 95%CI: 1.02–1.84), but not among women (AOR: 0.91, 95%CI: 0.67–1.23). No models among both men and women showed a significant association between short sleep and IDS. Long sleep duration was associated with PDS (AOR: 2.04, 95%CI: 1.20–3.48) among men but not among women (AOR: 1.26, 95%CI: 0.76–2.11). | Long but not short sleep duration was independently associated with IDS and PDS among men but not among women. |
| Tuo-Yu Chen et al, 2021 | 1/9/2021 | Cohort study. | We systemically investigated the temporal effects by examining the links between different insomnia symptom subtypes and the onset of depression at different follow-up intervals among community-dwelling older adults. | We used secondary data from the Health and Retirement Study (HRS). | The final sample size was 8080 for the analysis at the 2-year follow-up, 6407 for the analysis at the 4-year follow-up and 5347 for the analysis at the 6-year follow-up. | Insomnia symptom subtypes | Depression | 6 years. | Individuals with initial insomnia at baseline were about 33% more likely to develop depression at the 2-year (1.33[1.07-1.64]) and 4-year (1.33[1.01-1.74]) follow-ups. Late insomnia at baseline predicted the onset of depression at the 2-year follow-up (1.27[1.03-1.56]). Nonrestorative sleep predicted the incidence of depression at all follow-up periods (2-year: 1.36 [1.09-1.69]; 4-year: 1.33 [1.0001-1.76]; 6-year: 1.55 [1.12-2.15]). | This study finds that insomnia symptom subtypes may have different temporal effects on the future development of depression among community-dwelling older adults. |
| A. A. Kandola et al, 2021 | 6/17/2021 | Cohort study. | We conducted a prospective cohort study to determine how accelerometer-derived sedentary behaviour is associated with depression and anxiety symptoms while accounting for physical activity and sleep in a 24-h period and estimate the effect of replacing daily sedentary time with other movement behaviours (sleep, light, and moderate-to- vigorous activity) on the depression and anxiety symptoms. | Data from the UK Biobank. | 60,235 UK Biobank participants were included. | Sedentary behaviour, light activity, moderate-to-vigorous activity, and sleep. | Depression and anxiety symptoms | 4 years. | Replacing 60 min of sedentary behaviour with light activity, moderate-to-vigorous activity, and sleep was associated with lower depression symptom scores by 1.3% (95%CI, 0.4–2.1%), 12.5% (95%CI, 11.4–13.5%), and 7.6% (95%CI, 6.9–8.4%), and lower odds of possible depression by 0.95 (95%CI, 0.94–0.96), 0.75 (95%CI, 0.74–0.76), and 0.90 (95% CI, 0.90–0.91) at follow-up. | We found that daily sedentary behaviour time at baseline was positively associated with depression and anxiety symptom scores at follow up. Theoretically replacing periods of daily sedentary behaviour with light, moderate-to-vigorous activity, or sleep at baseline was associated with lower depressive symptom scores at follow-up. |
| **The relationship between sleep and functional disability** | | | | | | | | | | |
| Yen‑Han Lee et al., 2022 | 2/22/2022 | Cohort study. | This study examined the associations of activities of daily living (ADL) and instrumental activities of daily living (IADL) with changes in sleep-related measurements among Chinese older adults. | Data from the Chinese Longitudinal Healthy Longevity Survey. | The final study sample included a total of 42,417 observations. | ADL and IADL. | Sleep quality and sleep duraiton. | 9 years. | Older adults with more ADL (HR=0.96, 95%CI 0.95, 0.97; p<0.01) and IADL limitations (HR=0.94, 95%CI 0.93, 0.94; P<0.01) had a higher risk of having declining sleep quality over time (2005–2014). In the second model, we observed that older adults with more ADL (HR=0.98, 95%CI 0.96, 0.99; P<0.01) and IADL limitations (HR=0.96, 95%CI 0.96, 0.97; P<0.01) had a higher risk of transitioning from meeting the recommended sleep duration to not meeting the recommended sleep duration from 2005 to 2014. | Older adults with more ADL and IADL limitations had a higher risk of experiencing declines in sleep quality and the transition from meeting to not meeting the recommended sleep duration over time. |
| Mengli Liu et al., 2021 | 1/29/2021 | Cohort study. | To explore the effect of sleep duration at baseline on the incident IADL disability among middle-aged and older Chinese, and test whether cognition mediates this causality. | Data were collected from wave 1 (2011-2012) to wave 3 (2015-2016) of the China Health and Retirement Longitudinal Study (CHARLS). | 10,328 individuals were included in the study. | Sleep duration. | IADL disability. | 4 years. | Compared to 7-8 h sleep duration, both short sleep (OR=1.460; 95%CI: 1.261-1.690 for sleeping ≤5 h; OR=1.189; 95%CI: 1.011-1.400 for sleeping 5-7 h) and long sleep (OR=1.703; 95%CI: 1.269-2.286 for sleeping >9 h) were linked with incident IADL disability. KHB method identified significant mediating effect of cognition on the relationship between extreme sleep durations (≤5 h or >9 h) and IADL disability and the proportional mediation through cognition was 21.32% and 21.06% for sleeping ≤5 h and >9 h, respectively. | Compared to the sleep duration of 7-8 h, both short (sleep duration of ≤5 h and 5-7 h) and long sleep duration (sleep duration of >9 h) were predictors of IADL disability. And cognition partially mediated the effect of sleep duration on IADL disability. |
| Marcela Z. Campanini et al, 2018 | 6/11/2018 | Cohort study. | To examine the prospective association of sleep duration and quality with a range of measures of impairment in physical function and disability in older adults. | Data from participants in the Seniors-ENRICA (2012-2015, n=1,773) and in the ELSA cohort (waves 4 and 6, n=4,885) aged ≥60 years. | 6,658 individuals were included in the study. | Sleep duration and sleep quality. | Physical function impairment and disability. | 3-4 years. | Poor general sleep quality was linked to higher risk of impaired agility [OR: 1.93 (95% CI: 1.30-2.86) in Seniors-ENRICA and 1.65 (1.24-2.18) in ELSA study] and mobility [1.46 (0.98-2.17) in Seniors-ENRICA and 1.59 (1.18-2.15) in ELSA study]. Poor general sleep quality was also associated with decreased physical component summary (PCS) [1.39 (1.05-1.83)], disability in instrumental activities of daily living [1.59 (0.97-2.59)] and in basic activities of daily living [1.73 (1.14-2.64)] in Seniors-ENRICA. | We found that accumulation of sleep complains in older individuals independently predict physical function impairment and disability. |
| Shanshan Yang et al., 2023 | 11/4/2023 | Cohort study. | To comprehensively describe the characteristic of daytime, night and total sleep duration, sleep quality and diferent sleep mode of Hainan centenarians and their associations with activity of daily living (ADL) functions | Data based on the China Hainan Centenarians Cohort Study (CHCCS). | A total of 994 centenarians were included in this study. | Sleep duration and sleep quality. | ADL disability. | 2 years. | Compared with the centenarians who sleep 6–9 h at night and<2 h in the daytime, the adjusted OR between sleep>9 h at night and sleep≥2 h in the daytime and ADL disability was 2.93 (95%CI: 1.02–8.44), and adjusted OR of ADL mod erate & severe disability was 2.75 (95%CI: 1.56–4.83). Compared with centenarians who sleep for 7–9 h and have good sleep quality, centenarians who sleep for>9 h and have poor sleep quality have an increased risk of ADL moderate & severe disability (OR=3.72, 95%CI: 1.54–9.00). | Relation between sleep duration and ADL disability was more signifcant compared with sleep quality in Hainan centenarians. Poor sleep quality can aggravate the relationship between sleep duration and ADL moderate & severe disability. |
| Qing-Mei Huang et al., 2024 | 6/5/2024 | Cohort study. | To investigate the associations of insomnia symptoms and trajectories with functional disability. | Data from the Health and Retirement Study (HRS). | 13,197 participants were included. | Insomnia symptoms. | Functional disability. | 4 years. | Participants experiencing one (HR, 1.21; 95%CI, 1.13–1.29), two (HR, 1.43; 95%CI, 1.29–1.57), or three to four (HR, 1.41; 95%CI, 1.25–1.60) insomnia symptoms had a higher risk of ADL disability than asymptomatic respondents. Similarly, participants with one or more insomnia symptoms had a higher risk of IADL disability. Furthermore, using the trajectory with low insomnia symptoms as the reference, decreasing insomnia symptoms (HR, 1.22; 95%CI, 1.12–1.34), increasing insomnia symptoms (HR, 1.21; 95%CI, 1.05–1.41), and high insomnia symptoms (HR, 1.36; 95%CI, 1.18–1.56) were all associated with an increased risk of ADL disability. | Both a single measurement and dynamic trajectory of insomnia symptoms are associated with the onset of ADL disability. |
| **The relationship between sleep, depression and disability** | | | | | | | | | | |
| Jennifer L. Martin et al, 2010 | 5/7/2010 | Cohort study. | To describe sleep patterns in older adults living in assisted living facilities (ALFs) and to explore the relationship between sleep disturbance and quality of life, functional status, and depression over 6 months. | The study design was a prospective, observational cohort study in older people residing in a sample of ALFs located in the Los Angeles area. Participant recruitment was performed between April 2006 and March 2008. | One hundred twenty-one ALF residents. | Sleep parameters. | Quality of life, functional status, and depression. | 6 months. | Lower nighttime percentage sleep (t=2.65, P=0.009) was associated with worse baseline ADL scores. Lower percentage nighttime sleep predicted lower ADL functioning at 3 and 6 months, worse self-reported sleep quality, more nighttime awakenings (according to actigraphy), and high sleep apnea risk predicted more depressive symptoms at 3 months, and worse self-reported sleep quality and more nighttime awakenings predicted more depressive symptoms at 6 months. | Sleep disturbance is common in older ALF residents, and poor sleep is associated with declining functional status and quality of life and greater depression over 6 months of follow-up. |
| Tiina Paunio et al, 2015 | 10/14/2014 | Cohort study. | We aimed to examine association of self-reported sleep quality with incident symptoms of depression and disability retirement due to depressive disorders in a longitudinal population-based sample of twins. | Data from the Finnish Twin Cohort. | The sample included altogether 10,059 persons. | Sleep quality. | Depressive symptoms and disability pensions. | 15 years. | Onset of poor sleep between 1975 and 1981 predicted incident depression (BDItot OR=4.5, 95%CI:2.7–7.4, BDINATS OR=2.0, 95%CI: 1.4 – 2.7), while persistent poor sleep showed somewhat weaker effects (BDItot; OR=2.5, 95%CI:1.0–6.0, BDINATS OR=1.9, 95%CI:1.1–3.3). Among those with few recent stressful life events, onset of poor sleep predicted strongly depression (BDINATS OR=9.5, 95%CI:3.7–24.2). Likewise onset of poor sleep by 1981 increased the risk of disability retirement due to depression(OR=2.9, 95%CI: 1.8– 4.9) with a similar risk among those with persistent poor sleep (OR=2.7, 95%CI:1.3–5.7). | Poor sleep is an independent and robust risk factor for the various symptomatic domains of depression and for disability retirement due to depressive disorder. |

**Supplementary Table S3.** Characteristics of excluded studies.

| **Study** | **Reason for exclusion** |
| --- | --- |
| Lisa C. Barry et al., 2009 | Misalignment with the study aims |
| I. Carrie`re et al., 2009 | No research method |
| Jiska Cohen-Mansfield et al., 2010 | Misalignment with the study aims |
| Laura N. Gitlin et al., 2007 | Misalignment with the study aims |
| Banu ÖZYÜKSEL et al., 2007 | Wrong research subjects |
| Andrea Russo et al., 2007 | Wrong study design |
| Scott Schieman et al., 2007 | Misalignment with the study aims |
| Helen Street et al., 2007 | Misalignment with the study aims |
| Hana Vankova et al., 2008 | Wrong study design |
| Julie Hicks Patrick et al., 2004 | Wrong study design |
| Martha L Bruce, 2001 | Wrong study design |
| Kee-Lee Chou, 2007 | Misalignment with the study aims |
| Gerda M. Van der Weele et al., 2009 | Wrong study design |
| Haruna SAITO et al., 2008 | Misalignment with the study aims |
| Özlem Bozo et al., 2009 | Wrong study design |
| Aideen Freyne et al., 2005 | Misalignment with the study aims |
| Tameka Roberson BS et al., 2003 | Wrong research subjects |
| Shiau-Fang Chao, 2014 | Misalignment with the study aims |
| Zheng Ouyang et al., 2014 | Wrong study design |
| Noor Ani Ahmad et al., 2020 | Wrong study design |
| Bum Jung Kim et al., 2018 | Wrong study design |
| Lucienne A. Reichardt et al., 2020 | Wrong research subjects |
| Marzieh Mohamadzadeh et al., 2020 | Wrong study design |
| Sae Hwang Han et al., 2021 | Misalignment with the study aims |
| An Li et al., 2021 | Misalignment with the study aims |
| Maria Averina et al., 2005 | Misalignment with the study aims |
| Kimberly A. Babson et al., 2010 | Misalignment with the study aims |
| Chiara Baglioni et al., 2011 | Wrong study design |
| BJØRN BJORVATN et al., 2014 | Wrong study design |
| Bonanni E et al., 2010 | Misalignment with the study aims |
| Glenna S. Brewster et al., 2015 | Misalignment with the study aims |
| Daniel P. Chapman et al., 2013 | Wrong study design |
| Cheng-Sheng Chen et al., 2007 | Misalignment with the study aims |
| TSURUHEI SUKEGAWA et al., 2003 | Wrong study design |
| Daniel Foley et al., 2004 | Misalignment with the study aims |
| Daniel J. Foley et al., 2007 | Wrong study design |
| Won-Hyoung Kim et al., 2013 | Misalignment with the study aims |
| BERND KUNDERMANN et al., 2007 | Wrong research subjects |
| Pedro Ángel LATORRE-ROMÁN et al., 2015 | Misalignment with the study aims |
| Marie-France leblanc et al., 2015 (Clinical Interventions in Aging) | Wrong study design |
| Marie-France leblanc et al., 2015 (Nature and Science of Sleep) | Wrong study design |
| Markus Loeffler et al., 2015 | Misalignment with the study aims |
| Annemarie I. Luik et al., 2015 (DEPRESSION AND ANXIETY) | Wrong study design |
| Annemarie I. Luik et al., 2015 (J Sleep Res) | Misalignment with the study aims |
| Jianfeng Luo et al., 2013 | Misalignment with the study aims |
| Jeanne E. Maglione et al., 2012 | Wrong study design |
| J.E. McHugh et al., 2011 | Wrong study design |
| Ilona Merikanto et al., 2013 | Misalignment with the study aims |
| Sarosh J. Motivala et al., 2006 | Wrong study design |
| Maurice M. Ohayon et al., 2003 | Wrong study design |
| Paul Sadler et al., 2013 | Wrong study design |
| Stephen F. Smagula et al., 2013 | Wrong study design |
| Chia-Yi Wu et al., 2012 | Wrong study design |
| Yoram Barak et al., 2020 | Wrong study design |
| Nathália B. Becker et al., 2016 | Misalignment with the study aims |
| Anders Brostrom et al., 2017 | Wrong study design |
| Enda M. Byrne et al., 2019 | Wrong study design |
| Shun-Chiao Chang et al., 2016 | Misalignment with the study aims |
| Hung-Chun Lai et al., 2019 | Wrong study design |
| Yuning Liu et al., 2018 | Wrong study design |
| Tingting Sha et al., 2019 | Misalignment with the study aims |
| Christopher A. Webb et al., 2017 | Wrong study design |
| Kaitlin Hanley White et al., 2017 | Wrong study design |
| Chia-Rung Wu et al., 2019 | Wrong study design |
| Seongryu Bae et al., 2023 | Wrong study design |
| Katie Moraes de Almondes et al., 2022 | Misalignment with the study aims |
| Chaonan Du et al., 2024 | Misalignment with the study aims |
| RafaelGenario et al., 2023 | Wrong study design |
| Odessa S. Hamilton et al., 2023 | Misalignment with the study aims |
| Hanna K. Hausman et al., 2022 | Misalignment with the study aims |
| Min-Fang Hsu et al., 2021 | Wrong study design |
| Michael R. Irwin et al., 2021 | Wrong study design |
| Xiaowen Ji et al., 2020 | Wrong study design |
| Li Chunnan et al., 2021 | Wrong study design |
| Chien-Yu Lin et al., 2021 | Wrong study design |
| Haixia Liu et al., 2021 | Wrong study design |
| Huiying Liu et al., 2021 | Misalignment with the study aims |
| Leslie M. Swanson et al., 2023 | Wrong study design |
| Robert J. Zhou et al., 2023 | Misalignment with the study aims |
| Eric J. Lenze et al., 2005 | Wrong research subjects |
| Johan Ormel et al., 2002 | Wrong research subjects |
| Dimitris N. Kiosses et al., 2005 | Misalignment with the study aims |
| Namkee G. Choi et al., 2007 | Wrong research subjects |
| Julia D. Buckner et al., 2008 | Wrong study design |
| Simon Overland et al., 2008 | Outcome out of scope |
| Sally Wai-chi Chan et al., 2009 | Wrong research subjects |
| Jordan F. Karp et al., 2009 | Wrong research subjects |
| Erin Koffel et al., 2010 | Wrong study design |
| JULIEN MENDLEWICZ et al., 2009 | Misalignment with the study aims |
| Holly J. Ramsawh et al., 2008 | Wrong study design |
| Ryuji Furihata et al., 2011 | Wrong study design |
| Arjan W. Braam et al., 2014 | Misalignment with the study aims |
| Celia F. Hybels et al., 2015 | Wrong study design |
| Irene Romera et al., 2014 | Wrong research subjects |
| Kimberly A. Van Orden et al., 2016 | Wrong study design |
| Katarzyna B Malinowska et al., 2016 | Wrong study design |
| Myrthe C. Bruin et al., 2018 | Misalignment with the study aims |
| Daniel C. Mograbi et al., 2017 | Wrong study design |
| Lydia Poole et al., 2018 | Wrong study design |
| Le Xiao et al., 2018 | Wrong research subjects |
| Sonia Difrancesco et al., 2018 | Wrong research subjects |
| Junhyun Kwon et al., 2019 | Misalignment with the study aims |
| Agneta Lindegård et al., 2019 | Misalignment with the study aims |
| Cai W et al., 2020 | Misalignment with the study aims |
| Raquel Fábrega-Cuadros et al., 2020 | Misalignment with the study aims |
| Manqiong Yuan et al., 2019 | Misalignment with the study aims |
| Dao-min Zhu et al., 2020 | Wrong research subjects |
| Rebecca Chau et al., 2019 | Misalignment with the study aims |
| Komulainen K et al., 2021 | Misalignment with the study aims |
| Alexander Maier et al., 2021 | Misalignment with the study aims |
| Chao Wu et al., 2021 | Misalignment with the study aims |
| Rumei Yang et al., 2021 | Misalignment with the study aims |
| Jie Zhao et al., 2020 | Wrong research subjects |
| Alejandra Aguilar-Latorre et al., 2022 | Wrong study design |
| Salmaan Ansari et al., 2022 | Misalignment with the study aims |
| Antti Etholen et al., 2022 | Misalignment with the study aims |
| Yuanyuan Fu et al., 2020 | Misalignment with the study aims |
| Yingyun Hu et al., 2022 | Misalignment with the study aims |
| H. Ariel Bard et al., 2023 | Wrong study design |
| Lunan Gao et al., 2022 | Wrong study design |
| Dan Han et al., 2023 | Misalignment with the study aims |
| Shuang Han et al., 2023 | Misalignment with the study aims |
| Ching-Jow Hsieh et al., 2023 | Misalignment with the study aims |
| Soyoung Jang et al., 2023 | Misalignment with the study aims |
| Yunyi Wu et al., 2023 | Misalignment with the study aims |
| Manacy Pai et al., 2023 | Misalignment with the study aims |
| Ruijia You et al., 2023 | Misalignment with the study aims |
| Nima Javadzade et al., 2024 | Wrong study design |
| Xin Ye et al., 2024 | Misalignment with the study aims |
| Kexin Zhou et al., 2024 | Wrong study design |
| Yecun Liu et al., 2024 | Wrong study design |

**Supplementary Table S4.** Critical Appraisal Skills Programme Checklist (2024): For Cohort Studies.

| **Study ID** | **Critical Appraisal Skills Programme Checklist (2024): For Cohort Studies** | | | | | | | | | | | | | |
| --- | --- | --- | --- | --- | --- | --- | --- | --- | --- | --- | --- | --- | --- | --- |
|  | **Section A: Are the results valid?** | | | | | | | | **Section B: What are the results?** | | | **Section C: Will the results help locally?** | | |
|  | 1. Did the study address a clearly focused issue? | 2. Was the cohort recruited in an acceptable way? | 3. Was the exposure accurately measured to minimise bias? | 4. Was the outcome accurately measured to minimise bias? | 5. (a) Have the authors identified all important confounding factors? | 5. (b) Have they taken account of the confounding factors in the design and/or analysis? | 6. (a) Was the follow up of subjects complete enough? | 6. (b) Was the follow up of subjects long enough? | 7. What are the results of this study? | 8. How precise are the results? | 9. Do you believe the results? | 10.Can the results be applied to the local population? | 11.Do the results of this study fit with other available evidence? | 12.What are the implications of this study for practice? |
| **The relationship between depression and disability** | | | | | | | | | | | | | | |
| Kivela S.L. et al., 2001 | Yes | Yes | Yes | Yes | No | Yes | No | Yes | Yes | Yes | Yes | No | Yes | Yes |
| S.W.GEERLINGS et al., 2001 | Yes | Yes | Yes | Yes | No | Yes | No | Yes | Yes | Yes | Yes | Yes | Yes | Yes |
| Brenda W.J.H. Penninx et al., 2001 | Yes | Yes | Yes | Yes | Yes | Yes | Yes | Yes | Yes | Yes | Yes | Yes | Yes | Yes |
| Jingmei JIANG et al., 2002 | Yes | Yes | Yes | Yes | Yes | Yes | Yes | Yes | Yes | Yes | Yes | No | Yes | Yes |
| Li Wang et al., 2002 | Yes | Yes | No | Yes | Yes | Yes | Yes | Yes | Yes | Yes | Yes | No | Yes | Yes |
| Kala M. Mehta et al., 2002 | Yes | Yes | Yes | Yes | Yes | Yes | Yes | No | Yes | Yes | Yes | Yes | Yes | Yes |
| Johan Ormel et al., 2002 | Yes | Can’t Tell | Yes | Yes | No | No | No | No | Yes | Yes | Yes | No | Yes | Yes |
| Päivi Lampinen et al., 2003 | Yes | Can’t Tell | Yes | Yes | No | Yes | No | Yes | Yes | Yes | Yes | No | Yes | Yes |
| Jingmei Jiang et al., 2004 | Yes | Yes | Yes | Yes | Yes | Yes | Yes | Yes | Yes | Yes | Yes | No | Yes | Yes |
| SUSAN A. EVERSON -ROSE et al., 2005 | Yes | Yes | Yes | Yes | No | Yes | Yes | Yes | Yes | Yes | No | No | No | Yes |
| Nicholas P. Emptage et al., 2005 | Yes | Yes | Yes | Yes | Yes | Yes | Yes | Yes | Yes | Yes | Yes | No | Yes | Yes |
| Dorothy D. Dunlop et al., 2005 | Yes | Yes | Yes | Yes | Yes | Yes | Yes | No | Yes | Yes | Yes | Yes | Yes | Yes |
| Jason Schnittker, 2005 | Yes | Yes | Yes | Yes | No | Yes | Yes | Yes | Yes | Yes | Yes | Yes | Yes | Yes |
| YANG YANG et al., 2005 | Yes | Yes | Yes | Yes | Yes | Yes | Yes | Yes | Yes | No | Yes | Yes | Yes | Yes |
| Coen H. van Gool et al., 2005 | Yes | Yes | Yes | Yes | No | Yes | Yes | Yes | Yes | No | Yes | Yes | Yes | Yes |
| Eric J. Lenze et al, 2005 | Yes | Yes | Yes | Yes | No | Yes | Yes | Yes | Yes | Yes | Yes | Yes | Yes | Yes |
| F. Curtis Breslin et al., 2006 | Yes | Yes | Yes | Yes | No | Yes | Yes | Yes | Yes | Yes | Yes | Yes | Yes | Yes |
| Mathew D. Gayman et al., 2008 | Yes | Yes | Yes | Yes | No | Yes | Yes | Yes | Yes | No | Yes | No | No | Can’t Tell |
| Naoki Kondo et al., 2008 | No | Can’t Tell | Yes | Yes | No | Yes | No | No | Yes | Yes | Yes | No | Yes | Yes |
| Lydia W. Li et al, 2008 | Yes | Yes | Yes | Yes | No | Yes | Yes | Yes | Yes | Yes | Yes | Yes | Yes | Yes |
| Lisa C. Barry et al., 2009 | Yes | Yes | Yes | Yes | Yes | Yes | No | Yes | Yes | Yes | Yes | No | Yes | Yes |
| Milan Chang et al., 2009 | Yes | Yes | Yes | Yes | Yes | Yes | No | Yes | Yes | Yes | Yes | No | Yes | Yes |
| Kenneth E. Covinsky et al., 2010 | Yes | Yes | Yes | Yes | Yes | Yes | Yes | Yes | Yes | Yes | Yes | Yes | Yes | Yes |
| Celia F. Hybels et al., 2009 | Yes | Yes | Yes | Yes | Yes | Yes | Yes | Yes | Yes | Yes | Yes | Yes | Yes | Yes |
| Hajime Iwasa et al., 2009 | Yes | Yes | Yes | Yes | Yes | Yes | No | Yes | Yes | Yes | Yes | No | Yes | Yes |
| Carolyn L. Turvey et al., 2009 | Yes | Yes | No | Yes | No | Yes | Yes | No | No | No | Yes | Yes | Yes | Yes |
| Mark I. Weinberger et al., 2009 | Yes | Yes | Yes | Yes | No | Yes | No | No | Yes | Yes | Yes | No | Yes | Yes |
| Luca Dalle Carbonare et al., 2009 | Yes | Yes | Yes | Yes | Yes | Yes | Yes | Yes | Yes | Yes | Yes | Yes | Yes | Yes |
| Mari Kazama et al., 2011 | Yes | Can’t Tell | Yes | Yes | Yes | Yes | No | No | Yes | Yes | Yes | No | Yes | Yes |
| Erin Dunne et al., 2011 | Yes | No | Yes | Yes | No | Yes | No | Yes | No | No | No | No | Yes | Yes |
| Isabelle Carrière et al, 2011 | Yes | Yes | Yes | Yes | Yes | Yes | Yes | Yes | Yes | Yes | Yes | Yes | Yes | Yes |
| Chun-Min Chen et al., 2012 | Yes | Yes | Yes | Yes | Yes | Yes | No | Yes | Yes | Yes | Yes | No | Yes | Yes |
| Chun-Te Lee et al., 2012 | Yes | Yes | Yes | Yes | No | Yes | No | Yes | Yes | Yes | Yes | Yes | Yes | Yes |
| Xia Li et al, 2012 | Yes | Yes | Yes | Yes | No | Yes | No | Yes | Yes | Yes | Yes | No | Yes | Yes |
| Ya-Ting Yang et al., 2015 | Yes | Yes | Yes | Yes | No | Yes | Yes | Yes | Yes | Yes | Yes | No | Yes | Yes |
| Mary Elizabeth Bowen et al, 2015 | Yes | Yes | Yes | Yes | No | Yes | Yes | Yes | Yes | Yes | Yes | No | Can’t Tell | Yes |
| Kathryn L. Bacon et al., 2016 | Yes | Yes | Yes | Yes | No | Yes | No | Yes | Yes | Yes | Yes | No | Can’t Tell | Yes |
| Jin-Won Noh et al., 2016 | Yes | Yes | Yes | Yes | No | Yes | Yes | Yes | Yes | Yes | Yes | No | Can’t Tell | Yes |
| Takahiro Nakamura et al., 2017 | Yes | Can’t Tell | Yes | Yes | Yes | Yes | No | Yes | Yes | Yes | Yes | No | Yes | Yes |
| YANG YANG, 2016 | Yes | Yes | Yes | Yes | Yes | Yes | Yes | Yes | Yes | No | Yes | No | Yes | Yes |
| Kathrin Heser et al., 2018 | Yes | Yes | Yes | Yes | No | Yes | Yes | No | Yes | Yes | Yes | Yes | Yes | Yes |
| He Minfu et al., 2018 | Yes | Yes | Yes | Yes | Yes | Yes | Yes | No | Yes | Yes | Yes | Yes | Yes | Yes |
| Juliana Lustosa Torres et al., 2018 | Yes | Yes | Yes | Yes | Yes | Yes | Yes | Yes | Yes | Yes | Yes | Yes | Yes | Yes |
| Dexia Kong et al., 2019 | Yes | Can’t Tell | Yes | Yes | Yes | Yes | Yes | No | Yes | Yes | Yes | No | Yes | Yes |
| Ulrike Dapp et al., 2020 | Yes | Yes | Yes | Yes | No | Yes | Yes | Yes | Yes | Yes | Yes | No | Yes | Yes |
| Peter A. Coventry et al., 2020 | Yes | Yes | Yes | Yes | No | Yes | No | No | Yes | Yes | No | No | Yes | Yes |
| Rumei Yang et al., 2021 | Yes | Yes | Yes | Yes | Yes | Yes | Yes | Yes | Yes | Yes | Yes | Yes | Yes | Yes |
| Hongting Ning et al., 2021 | Yes | Yes | Yes | Yes | Yes | Yes | Yes | Yes | Yes | Yes | Yes | Yes | Yes | Yes |
| Fan Tian et al., 2022 | Yes | Yes | Yes | Yes | Yes | Yes | Yes | Yes | Yes | Yes | Yes | Yes | Yes | Yes |
| Mengxiao Hu et al., 2023 | Yes | Yes | Yes | Yes | Yes | Yes | Yes | Yes | Yes | Yes | Yes | Yes | Yes | Yes |
| Gina Lee et al., 2023 | Yes | Yes | Yes | Yes | No | Yes | No | Yes | Yes | Yes | Yes | No | Yes | Yes |
| Jiayi Wang et al., 2023 | Yes | Yes | Yes | Yes | Yes | Yes | Yes | Yes | Yes | Yes | Yes | Yes | Yes | Yes |
| Anda Botoseneanu et al., 2023 | Yes | Yes | Yes | Yes | No | Yes | Yes | Yes | Yes | Yes | Yes | Yes | Yes | Yes |
| Weihao Wang et al., 2024 | Yes | Yes | Yes | Yes | Yes | Yes | Yes | Yes | Yes | Yes | Yes | Yes | Yes | Yes |
| **The relationship between sleep and depression** | | | | | | | | | | | | | | |
| Lena Mallon et al., 2000 | Yes | Can’t Tell | Yes | Yes | No | Yes | Yes | Yes | Yes | Yes | Yes | No | Can’t Tell | Yes |
| Hyong Jin Cho et al., 2008 | Yes | Can’t Tell | Yes | Yes | Yes | Yes | No | No | Yes | Yes | Yes | No | Yes | Yes |
| Jae-Min Kim et al., 2009 | Yes | Yes | Yes | Yes | Yes | Yes | Yes | No | Yes | Yes | Yes | No | Yes | Yes |
| Isabelle Jaussen et al., 2011 | Yes | Yes | Yes | Yes | Yes | Yes | Yes | Yes | Yes | Yes | Yes | No | Yes | Yes |
| Misti Paudel et al., 2013 | Yes | Yes | Yes | Yes | Yes | Yes | Yes | Yes | Yes | Yes | Yes | No | Can’t Tell | Yes |
| Josine G. van Mill et al., 2013 | Yes | Yes | Yes | Yes | Yes | Yes | Yes | No | Yes | Yes | Yes | Yes | Can’t Tell | Yes |
| Jeanne E Maglione et al., 2014 | Yes | Yes | Yes | Yes | Yes | Yes | No | Yes | Yes | Yes | Yes | No | Yes | Yes |
| Lydia Poole et al., 2017 | Yes | Yes | Yes | Yes | Yes | Yes | Yes | Yes | Yes | Yes | Yes | Yes | Yes | Yes |
| Marta Jackowska et al., 2017 | Yes | Yes | Yes | Yes | Yes | Yes | Yes | Yes | Yes | Yes | Yes | Yes | Yes | Yes |
| Yujie Li et al., 2017 | Yes | Yes | Yes | Yes | Yes | Yes | Yes | No | Yes | Yes | Yes | Yes | Yes | Yes |
| Yankun Sun et al., 2018 | Yes | Yes | Yes | Yes | Yes | Yes | Yes | Yes | Yes | Yes | Yes | Yes | Yes | Yes |
| Michael J. Li et al., 2018 | Yes | Yes | Yes | Yes | No | Yes | No | No | Can’t Tell | No | Can’t Tell | No | Can’t Tell | Yes |
| Rize Jing et al., 2020 | Yes | Yes | Yes | Yes | Yes | Yes | Yes | Yes | Yes | Yes | Yes | Yes | Yes | Yes |
| Tuo-Yu Chen et al, 2021 | Yes | Yes | Yes | Yes | Yes | Yes | Yes | Yes | Yes | Yes | Yes | Yes | Yes | Yes |
| A. A. Kandola et al, 2021 | Yes | Yes | Yes | Yes | Yes | Yes | Yes | Yes | Yes | Yes | Yes | Yes | Yes | Yes |
| Annelies Brouwer et al., 2022 | Yes | Yes | Yes | Yes | No | Yes | Yes | Yes | Can’t Tell | Yes | No | No | No | Yes |
| Pengpid S et al, 2022 | Yes | Yes | Yes | Yes | Yes | Yes | Yes | Yes | Yes | Yes | Yes | Yes | Can’t Tell | Yes |
| Jialu Jiang et al., 2024 | Yes | Yes | Yes | Yes | Yes | Yes | Yes | Yes | Yes | Yes | Yes | Yes | Can’t Tell | Yes |
| **The relationship between sleep and disability** | | | | | | | | | | | | | | |
| Marcela Z. Campanini et al, 2018 | Yes | Yes | Yes | Yes | Yes | Yes | Yes | Yes | Yes | Yes | Yes | Yes | Yes | Yes |
| Mengli Liu et al., 2021 | Yes | Yes | Yes | Yes | Yes | Yes | Yes | Yes | Yes | Yes | Yes | Yes | Yes | Yes |
| Yen‑Han Lee et al., 2022 | Yes | Yes | Yes | Yes | No | Yes | Yes | Yes | Yes | Yes | Yes | Yes | Yes | Yes |
| Shanshan Yang et al., 2023 | Yes | Yes | Yes | Yes | Yes | Yes | No | No | Yes | Yes | Yes | No | Yes | Yes |
| Qing-Mei Huang et al., 2024 | Yes | Yes | Yes | Yes | Yes | Yes | Yes | Yes | Yes | Yes | Yes | Yes | Yes | Yes |
| **The relationship between sleep, depression and disability** | | | | | | | | | | | | | | |
| Jennifer L. Martin et al, 2010 | Yes | Can’t Tell | Yes | Yes | No | Yes | No | No | No | No | Can’t Tell | No | Yes | Yes |
| Tiina Paunio et al, 2015 | Yes | Yes | Yes | Yes | Yes | Yes | Yes | Yes | Yes | Yes | Yes | Can’t Tell | Yes | Yes |

**Supplementary Table S5.** Critical Appraisal Skills Programme Checklist (2024): Systematic Review.

| **Study ID** | **Critical Appraisal Skills Programme Checklist (2024): Systematic Review** | | | | | | | | | |
| --- | --- | --- | --- | --- | --- | --- | --- | --- | --- | --- |
|  | **Section A: Are the results of the review valid?** | | | | | **Section B: What are the results?** | | **Section C: Will the results help locally?** | | |
|  | 1. Did the review address a clearly focused question? | 2. Did the authors look for the right type of papers? | 3. Do you think all the important, relevant studies were included? | 4. Did the review’s authors do enough to assess quality of the included studies? | 5. If the results of the review have been combined, was it reasonable to do so? | 6. What are the overall results of the review? | 7. How precise are the results? | 8. Can the results be applied to the local population? | 9. Were all important outcomes considered? | 10. Are the benefits worth the harms and costs? |
| Jason E. Schillerstro et al., 2008 | Yes | Yes | No | Yes | Yes | Yes | No | Yes | Yes | Yes |
| Joyce T. Bromberger et al., 2009 | Yes | Yes | No | Yes | Yes | Yes | No | No | Yes | Yes |

**Supplementary Table S6.** Critical Appraisal Skills Programme Checklist (2024): For systematic reviews with meta-analysis of observational studies.

| **Study ID** | **Critical Appraisal Skills Programme Checklist (2024): For systematic reviews with meta-analysis of observational studies** | | | | | | | | | | | | | | | | | |
| --- | --- | --- | --- | --- | --- | --- | --- | --- | --- | --- | --- | --- | --- | --- | --- | --- | --- | --- |
|  | **Section A: Is the basic study design valid for a systematic review?** | | **Section B: Is the systematic review methodologically sound?** | | | | | | | **Section C: Are the results of the systematic review trustworthy?** | | | | | | | **Section D: Are the results of the systematic review relevant locally?** | **Section E: Will the implementation of the results represent greater value for your service users or population?** |
|  | 1. Did the systematic review address a clearly formulated research question? | 2. Did the researchers search for appropriate study design(s) to answer the research question? | 3. Were all the relevant primary research studies likely to have been included in the systematic review? a) Searching for primary research studies  b) Screening primary research studies from the search  c) Selecting primary research studies to include in the systematic review  d) Summarising the search and its outputs | | | | 4. Did the researchers assess the validity or methodological rigour of the primary research studies included in the systematic review? | 5. Did the researchers extract, and present information from the individual primary research studies appropriately and transparently? (a) Extraction of data  (b) Presentation of data | | 6. Did the researchers analyse the pooled results of the individual primary research studies appropriately?  6.1 Subgroup analysis  6.2 Meta-regression | | | 7. Did the researchers report any limitations of the systematic review and, if so, do the limitations discussed cover all the issues you have identified during critical appraisal?  7.1 Subgroup analysis  7.2 Meta-regression | | | 8. Would the benefits of acting upon the results outweigh any potential disadvantages, harms and/or additional demand for resources associated with acting on the results? | 9. Can the results of the systematic review be applied to your local population/in your local setting or context? | 10. If actioned, would the findings from the systematic review represent greater or additional value for the individuals or populations for whom you are responsible? |
| Xin-lin Li et al., 2023 | Yes | Yes | Yes | Yes | Yes | Yes | Yes | Yes | Yes | Yes | No | Yes | Yes | No | Yes | Can’t Tell | Yes | Yes |
